# Public health impact of current and proposed age-expanded perennial malaria chemoprevention: a modelling study

**DOI:** 10.1101/2024.07.31.24311277

**Authors:** Swapnoleena Sen, Lydia Braunack-Mayer, Sherrie L Kelly, Thiery Masserey, Josephine Malinga, Joerg J Moehrle, Melissa A Penny

**Affiliations:** Swiss Tropical and Public Health Institute, Allschwil, Switzerland; University of Basel, Basel, Switzerland; Medicines for Malaria Venture, Geneva, Switzerland; Telethon Kids Institute, Nedlands, WA, Australia; Centre for Child Health Research, The University of Western Australia, Crawley, WA, Australia

**Author notes:** Correspondence to: Prof Melissa A Penny.

## Abstract

In 2022, the World Health Organization extended their guidelines for perennial malaria chemoprevention (PMC) from infants to children up to 24 months old. However, evidence for PMC’s public health impact is primarily limited to children under 15 months. Further research is needed to assess the public health impact and cost-effectiveness of PMC, and the added benefit of further age-expansion. We integrated an individual-based model of malaria with pharmacological models of drug action to address these questions for PMC and a proposed age-expanded schedule (PMC+, for children 03-36 months). Across prevalence settings of 5-70% and different drug sensitivity assumptions, we predicted PMC and PMC+’s median efficacy of 18.6%(12.2-25.0%) and 21.9%(14.3-29.5%) against clinical disease and 9.0%(2.0-16.0%) and 10.8%(3.2-18.4%) against severe malaria, respectively, in children under three years. PMC’s total impact outweighed risk of delayed malaria in children up to age five and remained cost-effective when delivered through the Expanded Program on Immunization.

## Main

The World Health Organization (WHO) recommends perennial malaria chemoprevention (PMC) to protect children up to 24 months of age from *Plasmodium falciparum* malaria in settings with perennial transmission, and medium to high parasite prevalence greater than 10% among 2-10-year-olds. A course of prophylactic treatment with sulphadoxine-pyrimethamine (SP) is given at pre-specified ages through the Expanded Program on Immunization (EPI), regardless of a child’s infection status. The 2022 WHO guidelines [1] substantially revised previous recommendations for intermittent preventive treatment in infants (IPTi) to encourage broader adoption of this safe, affordable, yet underutilized malaria control intervention [1–7]. Multiple factors were thought to have contributed to the insufficient uptake of IPTi: inconsistent and relatively short duration of protection [8, 9], insignificant impact on mortality [10], a fixed dosing schedule (while EPI schedules vary by country), unclear eligibility of target seasonality settings, and deployment based on parasite genetic biomarkers (while many countries do not have regular genetic surveillance) [11]. With these major revisions, the WHO encourages PMC to be timed according to a country’s EPI schedule [12, 13]. The restriction to deploy it according to parasite genetic biomarker prevalence was also lifted, based on the accumulated evidence of SP’s maintained effectiveness against partially resistant parasites (such as, mutations in the *Pfdhfr* gene encoding dihydrofolate reductase and *Pfdhps* gene encoding dihydropteroate synthase [1, 14]).

There is currently limited information on the public health impact of the updated PMC [1]. Three clinical studies evaluated age-targeted SP’s efficacy in children up to 15 months [8, 15, 16], and a recent modelling study assessed it up to18 months of age [17]. Two more clinical trials investigated the program in children up to 24 months, but implemented different monthly dosing schedules than current PMC [9, 18]. Ongoing Plus Projects [12, 13] are important for co-designing contextual dosing for children under two years. However, no study has yet explored the potential for enhancing public health impact by incorporating further age-expansion into next-step designs [1].

While PMC is usually perceived to be safe, the risk of delayed malaria is often a concern. This is because a time-limited intervention that targets young children may interfere with the acquisition of natural immunity [19, 20]. To note, post-intervention effects are often referred interchangeably as rebound, leading to vague interpretations and an unclear understanding of the issue [19, 20]. The WHO’s Global Malaria Programme recently laid out a framework for systematically assessing post-intervention effects. However, there remains limited information of this risk due to the limited uptake of the revised PMC as yet [21], and early IPTi trials showed contradictory effects [2, 22, 23]. As a result, there is a need to collate evidence of PMC’s positive net benefit over the intervention and post-intervention period against clinical and severe malaria to support its wider deployment [19, 20].

In this context, mathematical modelling offers useful tools to assess the public health impact of malaria chemoprevention scale-up and policy-expansion [19, 21, 24, 25]. However, we found only one modelling study that assessed delayed malaria risk of PMC in children older than 12 months of age, and this study applied a monthly dosing schedule that differs from current PMC dosing recommendations [26]. Similarly, an economic analysis of PMC-SP in children aged 3 to 59 months was based on different dosing schedules [3]. Since additional EPI touch points exist up to 36 months of age (such as for booster doses of diphtheria and tetanus vaccines, deworming between 12–23 months, the second dose of the measles vaccine at 24 months, vitamin A supplementation in 6-59 months), reaching children up to 36 months could pave the way to increasing PMC’s impact through an age-expansion strategy [1, 6, 27].

In this study, we modelled two age-targeted SP dosing schedules (PMC for children 03-24 months, and PMC+ for 03-36 months of age) under various assumptions for drug resistance and chemoprevention coverage. These schedules were selected according to age-patterns of severe malaria risk and likely routine EPI touchpoints [1, 6, 12, 20]. We further compared our results to those using former IPTi deployment to support calibration of our drug model. We examined post-intervention effects in children up to age five, and explored possible mitigation strategies for any potential of delayed malaria [19, 20]. The cost-effectiveness for EPI-linked delivery of both PMC and PMC+ was assessed in various access to healthcare settings to inform potential implementation studies [28].

We evaluated the public health impact and cost-effectiveness of PMC and PMC+ in archetypal perennial and exemplar sub-perennial transmission settings (where malaria incidence increases during rainy seasons in addition to being present year-round [29]). These include all regions where PMC guidelines apply. Besides, hotspot perennial malaria prevalence is also found in countries experiencing primarily seasonal transmission (e.g., in Kenya [30]). Many of these areas fall outside the settings targeted by both perennial and seasonal malaria chemoprevention (SMC), but will likely benefit from a sustained PMC or PMC+ rollout (hereafter referred as “PMC(+)” to indicate either PMC or PMC+ deployment) [29, 31]. We aim to use this quantitative evidence to inform pilot implementation studies and new malaria chemoprevention guidelines to achieve a greater impact and reduce missed opportunities, particularly among the most vulnerable young children.

## Results

### Impact on malaria morbidity

PMC with seven SP doses and PMC+ with nine SP doses were simulated as continuous programs aligned with exemplar EPI touchpoints. To assess the public health benefits attained by PMC(+) and to support model calibration of SP, former IPTi was simulated with three SP doses in infants. All results were reported five years after the intervention rollout. Thereby, participants who were enrolled in the first year of the program had received all PMC(+) doses and reached post-intervention ages at the time of analysis.

Our results suggested that an age-expanded PMC schedule will likely improve impact in different transmission, parasite genotype, and clinical settings across perennial and sub-perennial seasonality (**Fig. 1a**). Overall, we predicted PMC and PMC+ to have median (interquartile range) efficacy of 18.6%(12.2-25.0%) and 21.9%(14.3-29.5%) against clinical malaria in children under three years, and 9.0%(2.0-16.0%) and 10.8%(3.2-18.4%) against severe malaria, respectively. Both PMC and PMC+ showed substantially increased efficacy compared to IPTi across settings (relative increase of efficacy of 118.3% and 152.65% against clinical and 62.95% and 97.05% against severe disease, respectively). We predicted increasing protection against clinical cases with increasing transmission, lower access to case management (first-line treatment with artemether-lumefantrine), and increasing program coverage. Consistent with earlier studies, our results indicated that SP remains largely effective as a chemoprevention drug in the face of partial SP resistance (such as, quadruple dhfr-51I, dhfr-59A, dhfr-108A, and dhps-437G mutations in *Pf*dhfr and *Pf*dhps genes) [12, 32–34]. Our results also suggest that increased efficacy by PMC’s age-expansion could potentially counter partial resistance and maintain desired levels of protection in children under three years. The increase in efficacy by PMC+ compared to PMC remained largely sustainable against clinical malaria in both partially SP-resistant and SP-sensitive settings (**Fig. 1b**). The efficacy increased modestly, but reduced uncertainty of benefit against severe malaria in SP-sensitive settings. As such, PMC+ was found to better cover children throughout the immunologically vulnerable ages [35]. However, any implications for the spread of drug resistance due to dose expansion should be interpreted with caution, as we will discuss later.

**Fig. 1.**
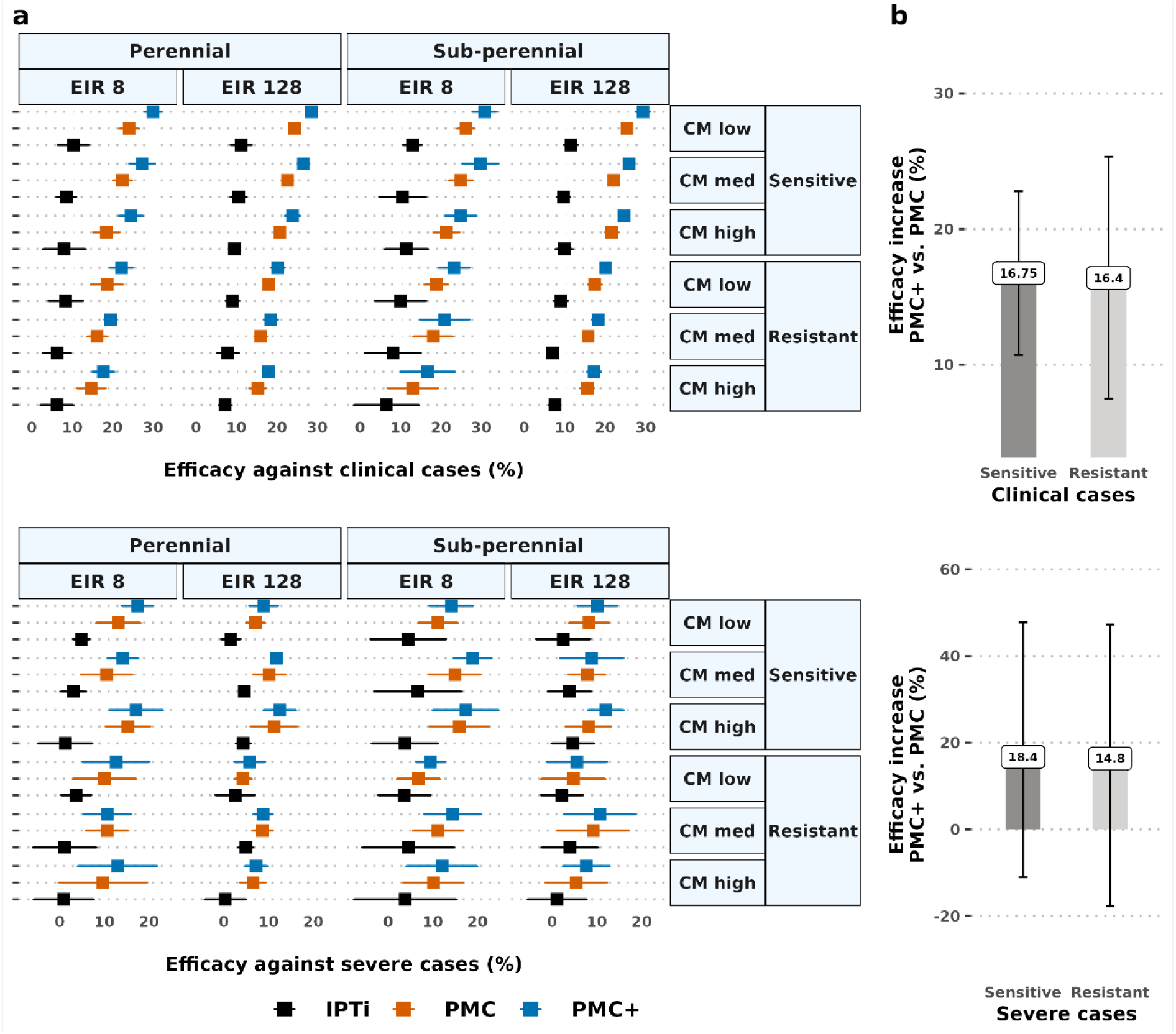
Protective efficacy against all episodes of clinical and severe malaria over the first three years of life, expressed as a relative reduction of the incidence rate ratio. The left plot in panel (a) shows the median efficacy and associated interquartile range in perennial and sub-perennial settings with medium (*Pf*PR_2-10_ 10-19%, EIR 8) to very high intensity of transmission (*Pf*PR_2-10_ 60-70%, EIR 128). Results depict the impact of dosing schedules for IPTi, PMC, and PMC+ simulated under full chemoprevention coverage (100% at each dosing cycle) in either SP-resistant (partial resistance conferred by *Pfdhfr* and *Pfdhps* mutant genotypes) or SP-sensitive (wild type *P. falciparum*) settings. The prophylactic period lasted 35 days in resistant settings compared to 42 days in sensitive settings. Varying healthcare strength is represented by low (10%), medium (30%), and high (50%) probability of accessing case management within 14-days post-diagnosis. The right plot in panel (b) depicts the relative increase of median efficacy (interquartile range) by PMC+ compared to PMC. CM: case management; EIR: entomological inoculation rate per person per year; IPTi: intermittent preventive treatment in infants; PMC: perennial malaria chemoprevention; PMC+: proposed age-expanded perennial malaria chemoprevention; *Pf*PR_2-10_: *P. falciparum* prevalence in 2-10-year-olds; SP: sulphadoxine-pyrimethamine.

As expected, PMC(+)’s total program coverage influenced its effectiveness. A coverage of 80% at each dosing cycle led to only 20% of children receiving all seven PMC doses, and only 10% of children receiving all nine PMC+ doses. This, in turn, led to differential impact against clinical and severe cases (**extended data Fig. E1a**). Although effectiveness against clinical cases consistently increased with higher chemoprevention coverage across settings, effectiveness against severe cases varied depending on both access to case management and assumptions about drug sensitivity (**extended data Fig. E1a and Fig. E1b**).

Furthermore, we tracked the cumulative cases that could be averted by expanding the age range of PMC. These findings could be used to advocate for benefits among implementation partners [12]. For instance, PMC+ reduced about 500 more clinical cases compared to PMC by the 5^th^ year post-deployment in settings with partial drug resistance in a village level population of 10,000 people (*Pf*PR_2-10_ 50-59%) (**extended data Fig. E2**).

### Post-intervention delayed malaria

For malaria chemoprevention targeting young children, it is important to monitor post-intervention effects. Although, based on SP’s shorter duration of protection, PMC with SP is less likely to interfere with the development of antimalarial immunity acquisition than other, longer acting or more efficacious chemoprevention [26]. We recorded a low risk of an age-shifted burden of malaria in children up to five years across the wide transmission range (**Fig. 2a**, **Fig. 2b**). While there is a shift, the net impact remains positive as is discussed below. The extent of delayed malaria was lower in a fully drug sensitive settings. Our results argue in favor of age-expansion to protect children against malaria throughout their most vulnerable years, since a low risk of delayed malaria was also predicted for PMC+. Children remained protected for up to six months after the last dose, despite SP having a relatively short period of protection. The age pattern of incidence was similar across settings with high levels of access to care for all modelled prevalence levels.

**Fig. 2.**
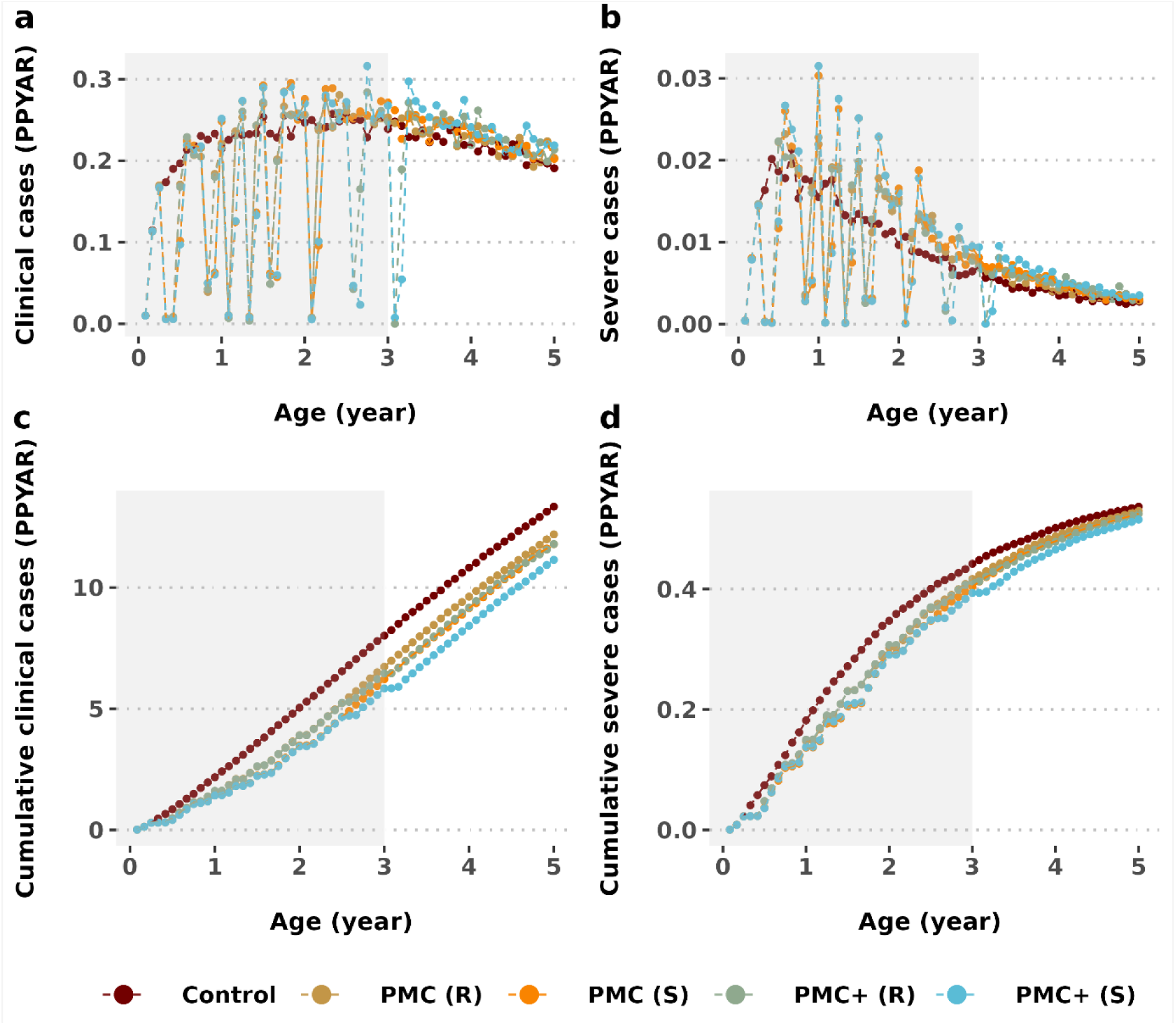
Post-intervention effects by age. The top panels (a, b) show the impact of PMC or PMC+ on the age pattern of malaria incidence in children under five compared to control group in perennial malaria transmission areas. The bottom panels (c, d) show the total impact, expressed as cumulative cases per age. Grey shaded areas depict the age targeted by each intervention. Results are depicted for settings with high prevalence (*Pf*PR_2-10_ 40-49%, EIR 32), medium access to treatment (30% probability within 14-days post-infection) and full chemoprevention coverage. PMC: perennial malaria chemoprevention; PMC+: proposed age-expanded perennial malaria chemoprevention; R: partially SP-resistant, S: fully SP-sensitive; *Pf*PR_2-10_ : *P. falciparum* prevalence in 2-10-year-olds; SP: sulphadoxine-pyrimethamine.

### Overall net intervention impact

A positive net benefit (i.e., total intervention and post-intervention impact) [19] of PMC(+) was sustainable in children up to five years (**Fig. 2c**, **Fig. 2d**). This net benefit increased in higher prevalence settings, outweighing the increased risk of delayed malaria. This was noted for both clinical and severe cases, especially when higher access to treatment was available. The positive net benefit against clinical cases was maintained for up to one year following the final dose of PMC, despite SP’s short period of protection.

The risk of delayed malaria and the extent of the program’s net benefit was driven by the characteristics of the setting, particularly for severe malaria [6] (**Fig. 3**). As anticipated, protection against severe malaria was largely influenced by the level of access to first-line treatment due to the need to counter age-shifted malaria cases. Our results indicate that in settings with moderate to high access to malaria treatment PMC was able to mitigate the risk of both delayed clinical and severe malaria. This emphasizes the need for strong healthcare systems, especially for adequate management of severe malaria cases once children are no longer eligible for PMC(+). Additionally, levels of baseline annual malaria prevalence were influenced by the level of access to first-line treatment for malaria cases (**extended data Fig. E3**). This reinforces the WHO’s ongoing emphasis on strengthening case management [36].

**Fig. 3.**
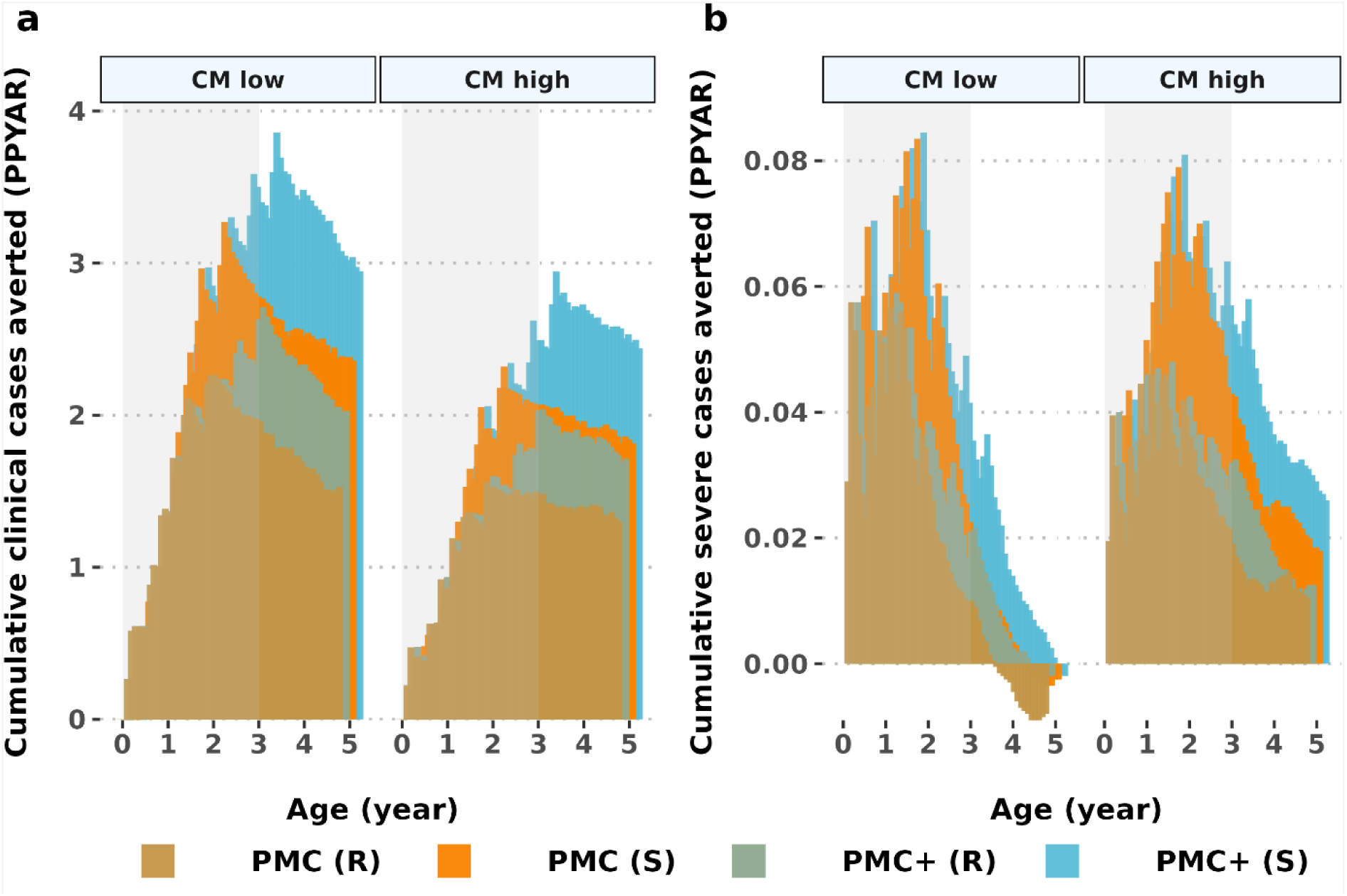
Cumulative cases averted in the intervention and follow-up ages under different levels of healthcare strength settings. The strength of healthcare systems is depicted by a low (10%) and high (50%) probability of having access to case management (within 14-days post-infection). Results are shown for settings with high prevalence (*Pf*PR_2-10_ 60-70%) in regions with low and high access to treatment and 100% chemoprevention coverage. Grey shaded areas indicate the period of intervention up to three years of age. CM: case management; PMC: perennial malaria chemoprevention; PMC+: proposed age-expanded perennial malaria chemoprevention; *Pf*PR_2-10_: *P. falciparum* prevalence in 2-10-year-olds; SP: sulphadoxine-pyrimethamine.

### Trends of cost-effectiveness

We assessed affordability in terms of incremental cost-effectiveness (ICER) of PMC(+) against malaria morbidity to inform decisions for possible implementation [1]. Given that SP is a low-cost drug, especially when delivered through the EPI, ICER was found to be aligned with the trends we observed above for PMC(+)’s effectiveness and total impact against clinical and severe cases. We anticipate affordability to be more favourable in higher transmission settings with lower access to treatment. The median ICER of either PMC or PMC+, measured in comparison to no chemoprevention, ranged between US$0.15 to US$0.8750 per clinical case averted (**Fig. 4a**), and between US$8.78 to US$22.88 per severe case averted (**extended data Fig. E4**) across WHO recommended transmission levels As a secondary economic analysis, we examined the total intervention costs, after accounting for treatment expenses for managing clinical malaria cases by using a first-line treatment artemether-lumefantrine (**Fig. 4b**). We found estimated cost savings across these settings between US$0.43 to US$1.15 per clinical case averted.

**Fig. 4.**
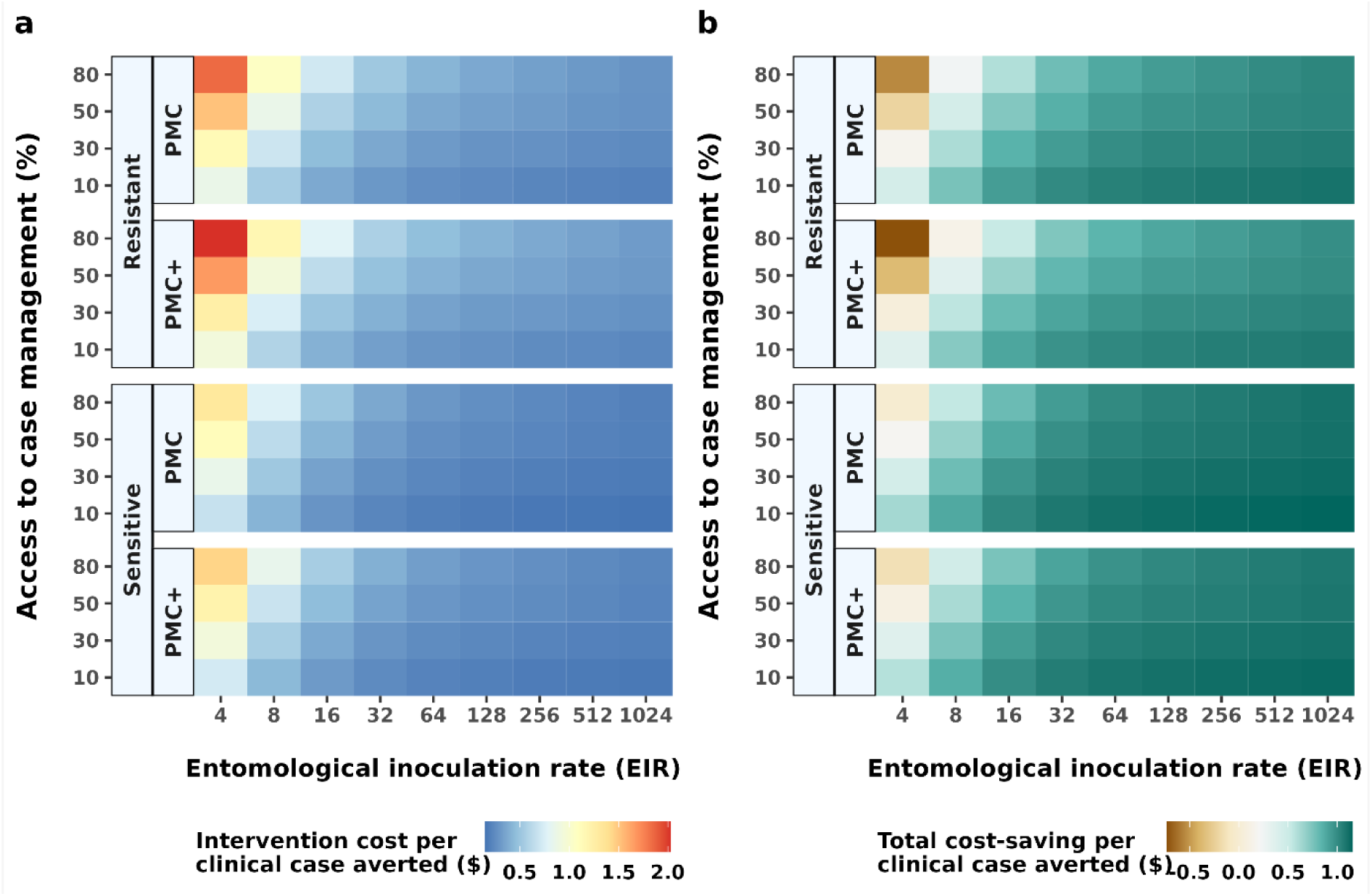
**Incremental cost effectiveness for PMC and PMC+ in partially SP-resistant and fully SP sensitive settings with various levels of access to treatment, compared to a counterfactual of no chemoprevention. The left panel (a) depicts the cost of PMC(+) delivery, while the right panel shows total cost-savings for treating clinical cases using first-line treatment with artemether-lumefantrine after accounting for PMC(+) delivery cost.** CM: case management; PMC: perennial malaria chemoprevention; PMC+: proposed age-expanded perennial malaria chemoprevention; SP: sulphadoxine-pyrimethamine.

Accordingly, the incremental cost-effectiveness of age-expanded PMC+ compared to current PMC (ICER_PMC+/PMC_) indicated higher cost-effectiveness in moderate to high transmission intensity settings (*Pf*PR_2-10_ >10%, EIR>16) for all levels of access to care (**Fig. 5a**). The net cost-effectiveness analysis included a combination of chemoprevention delivery and treatment cost-savings across different coverage levels and drug susceptibility scenarios (**Fig. 5b**). These values were determined by both the epidemiological setting characteristics and intervention coverage. Our results demonstrated better cost savings in SP-sensitive settings, even when PMC(+)’s coverage was reduced. On the contrary, the total cost increased under reduced coverage levels in resistant settings as compared to PMC with full coverage. In such settings, fewer clinical cases were estimated to have been averted, and thus higher total treatment costs were calculated. Our analysis assumed higher treatment cost per episode of clinical malaria (with artemether-lumefantrine) than per dose PMC(+) with SP delivery at their 2024 cost values (**supplementary Table S5**).

**Fig. 5.**
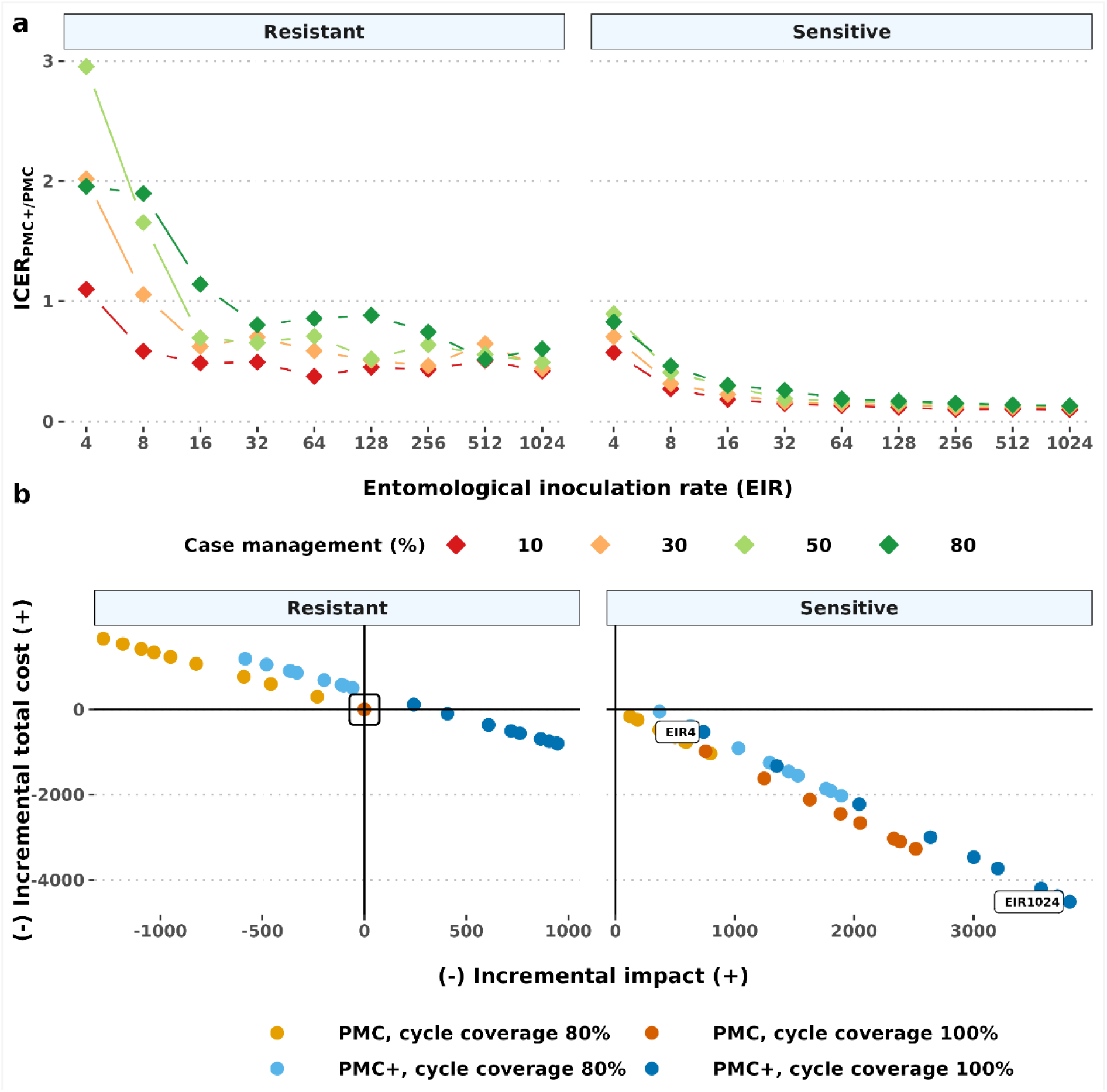
Projected incremental cost-effectiveness of PMC+ against clinical malaria compared to PMC (ICER_PMC+/PMC_) for a cohort of 1000 children. The top panel (a) denotes ICER_PMC+/PMC_ in different transmission (shown by entomological inoculation rates) and coverage levels. The bottom panel (b) depicts the total cost after accounting for treatment cost savings versus the number of clinical cases averted. The total cost was estimated by subtracting treatment cost from chemoprevention cost. The comparator is PMC with 100% coverage for all dosing cycles, in partial SP-resistant settings (indicated by the black box in panel b). Each point represents the total cost-effectiveness dynamics at each simulated entomological inoculate rate (EIR) relative to the respective comparator values in the same EIR. The cost-effectiveness benefit relative to the comparator was determined by the transmission and coverage levels. The trend of this is shown for PMC+ with 100% coverage in the sensitive setting by annotating the values at the lowest (EIR 4) and highest simulated entomological inoculation rate (EIR 1024). Results depict perennial settings. PMC: perennial malaria chemoprevention; PMC+: proposed age-expanded perennial malaria chemoprevention; SP: sulphadoxine-pyrimethamine.

### Validation of results against empirical data

Before collating these model-based insights, we validated our model’s (OpenMalaria) estimated effect sizes against an early randomized controlled IPTi trial [37]. In addition, all outputs were compared to a recent meta-analysis [2]. Another objective of comparing in-silico trial results to empirical data was to examine SP’s parasite life-stage specific mode of actions that included: either (1) a blood-stage action that clears existing infections or (2) a liver-stage prophylactic action that prevents new infections, or (3) a combined blood- and liver-stage action, assuming each effect is independent [ 38, 39]. The median protective efficacy (PE) of SP was evaluated by administering three doses to children at three, four, and nine months of age (**supplementary Table S2)**. Our results showed that combined blood- and liver-stage activity [40] overestimated the effect size (**extended data Fig. E5**). The range of PE from either only blood- or liver-stage activity corresponded to the empirical data range. However, we selected the blood stage parasite clearing action of SP for all simulations in this study. This is because existing evidence suggests that longer acting sulphadoxine is likely to have asexual blood-stage parasites clearance action, which drives the duration of prophylaxis [4, 28, 41–43]. As yet, there is no clear understanding of its liver stage activity [44–47]. Only a few pre-clinical studies revealed that the partner drug pyrimethamine may act against liver-stage parasites in animal models [44]. Altogether, blood stage clearance activity was applied to model all scenarios in this study. This way our model estimated effect size (median PE of 22.7% against clinical cases) closely matched to the empirical data (22.6% in the replicated IPTi trial [37] and 22% in the meta-analysis [2] (**supplementary Table S2 and Table S3**)). Additionally, consistent with the findings from the replicated trial, we found that PE from both the intention-to-treat schedule (that included all participants who received at least one SP dose) and per-protocol schedule (that included only participants receiving all doses) showed very low difference (**extended data Fig. E6**). All results presented in this study were performed following an intention-to-treat schedule. The protection lasted for about one month after each dose in partially SP-resistant settings, [39, 48], and the risk of delayed malaria was low [37] (**extended data Fig. E7**).

## Discussion

Our modeling study addressed important research needs to better understand the potential public health impact of the WHO’s revised (2022) perennial malaria chemoprevention (PMC) recommendations. We focused on generating quantitative evidence for the effectiveness and cost-effectiveness of PMC across recommended settings, alongside assessing potential post-intervention effects. We additionally examined a potential age-expanded delivery strategy (PMC+) for children up to 36 months of age. Our proposed deployment of PMC(+) leveraged existing routine EPI delivery channels [1, 31]. Although the current guidelines do not necessitate resistance biomarker-based deployment, we compared all outcomes in partially SP-resistant (quadruple mutation in *Pfdhfr* and *Pfdhps* genes) and fully SP-sensitive parasite settings. These results can inform discussions around the rationale of continued SP use across different parasite genotype settings. Overall, our study provides evidence for multiple open research questions to inform implementation decisions around the updated PMC recommendations. Furthermore, using a validated disease and intervention model, we estimated that expanding PMC to older children will reduce missed opportunities to prevent malaria during the most vulnerable ages.

We found that adding only two additional dosing cycles to PMC improved the intervention’s effectiveness by a median of 16.7% against clinical and 17.4% against severe malaria compared to the current PMC across the full range of prevalence, healthcare strength, drug sensitivity and seasonality settings. If more doses could be added through further EPI contacts or alternative delivery channels, the effectiveness will likely improve. Both PMC(+) schedules were shown to be more protective against clinical malaria in weaker healthcare systems. PMC+ achieves higher effectiveness compared to PMC against clinical malaria under reduced coverage as well (for instance, 80% coverage at each dosing cycle for PMC+ compared to higher coverage for each cycle of PMC). This will likely alleviate concerns against gradually reducing coverages and total impact for expanding number of chemoprevention doses.

We modelled varying assumptions concerning drug sensitivity utilising different half-maximal effective concentration (EC50) values [39]. In accordance with earlier findings, we found that SP’s chemopreventive effectiveness was largely maintained in settings with partially resistant quadruple mutant genotypes (where SP remains protective for about 35 days) [1, 49, 50]. Our results support earlier evidence showing that resistance conferring mutation is only one of the many factors that contribute to the chemoprevention effectiveness of a drug, including coverage, adherence, policy uptake, and the nutritional and immune status of recipients. [32]. Consistent with SMC modelling results [39], we speculate that the proposed age-expansion is unlikely to contribute to the spread of SP-resistance, as PMC+ is to be given to a comparable age group to PMC. However, the analysis of genetic biomarker survey data can better address such concerns in empirical settings. Specifically, monitoring the type of mutation is important, as higher prevalence of the quintuple mutant genotype (where SP’s prophylactic effect lasts for about 20 days [32]) could potentially further diminish chemoprevention efficacy.

This is the first study to evaluate the post-intervention effects for revised PMC recommendations, following the WHO’s guidelines and evaluating effects up to age five [19, 20]. Our understanding of PMC’s rebound dynamics is still inconclusive, due to the relatively short follow-up duration of one year after the last PMC dose, and due to the challenges of performing empirical studies with long follow-up [22, 23]. We did not find any trial that monitored these effects beyond age three. In this study, we monitored the age-pattern of incidence up to age five [19, 21]. Our results indicated low risk of age-shifting for both the “leaky” PMC and PMC+ deployment, consistent with the conclusions from the WHO’s report on rebound phenomenon [19]. However, patterns of risk differ between clinical and severe malaria. Higher net impact against severe malaria was recorded in settings with high access to case management, in contrast to higher impact against clinical cases in settings with low access to case management setting. These results suggest that the low risk of delayed severe malaria can be mitigated by strengthening health systems. PMC(+)’s total impact was shown to be higher in fully drug sensitive settings. This strengthens the rationale for continuing efforts to contain the spread of resistance and to explore alternative drug candidates, even though SP remains effective as a chemoprevention drug in partially resistant settings.

Consistent with other modelling studies [3], we found that PMC(+) will remain cost-effective in moderate to high transmission settings. Median ICER showed both PMC and PMC+ will likely remain affordable at under US$1 per clinical episode and under US$23 per severe episode of a potentially life threatening disease averted, across recommended levels of malaria transmissions (*Pf*PR_2-10_ above 10%, EIR 16 or more). Implementing PMC(+) resulted in cost savings up to US$1.15 per clinical case averted. PMC+ will likely be more affordable in regions with a higher intensity of transmission, drug-sensitivity, and lower access to case management. This result makes a strong case that PMC(+) will continue to deploy SP, given it is relatively cheap drug, and can be delivered through existing EPI [1, 12]. Moreover, increased cost savings was observed also under reduced coverage assumptions in SP-sensitive settings. These results indicated PMC(+) is likely to be substantially more affordable compared to other malaria chemoprevention, such as vaccines. For instance, median ICER for deploying RTS,S vaccine is predicted to be between $204 to $279 per clinical case averted [51].To note, estimates of detailed health savings and cost-effectiveness based on country data are beyond the scope of this work. Hence, we did not select any threshold of cost-effectiveness [52]. To realize health benefits in areas of reduced SP sensitivity, repurposed next-generation tools are needed that target resistant parasites. Also, to maintain a comparable health economic benefit, the cost of goods for these tools must closely align with those of SP. The possibility to deploy PMC(+) through channels alternative to EPI also remains open for future studies [3, 24, 53]. However, our findings indicate a positive trend of incremental cost-effectiveness for an age-expanded dosing schedule, providing valuable insights for pilot implementation studies.

To reliably predict a drug’s impact, it is important to understand its mode of action. Surprisingly, despite long and widespread use of SP for chemoprevention, there is no conclusive evidence for the combination drug’s parasite life-stage specific mode of action [42, 44]. Two modeling studies explored differing hypothesizes [4, 54]. Our model validation results suggest that SP blood-stage parasite clearing activity is the relevant mechanism of action, as aligned with earlier studies [41, 42], and this model was applied for all predictive analysis in this work.

As with any modeling study, there are several limitations to our results. First, as previously elaborated, our predictions are based on the dominant blood-stage activity of sulphadoxine, but clear evidence of SP as a combination may need further investigation. Our broad conclusions regarding the impact of PMC(+) will likely hold true, as shown in earlier studies that compared different modes of SP’s action [55]. However, this study is not suited to be able to conclude whether alternative modes of action (such as, only liver-stage prophylactic action or a combination of blood- and liver-stage activity) lead to differences in immunity acquisition. Second, it may be beneficial to expand PMC to children up to five years old, who remain at risk of malaria [56]. However, this will require alternative and reliable delivery channels as EPI touch points are not available for this age group. Currently, there is limited operational experience with such delivery options and no clear estimations of the cost for establishing and delivering through alternative channels [36]. Third, our model does not explicitly assess the impact of chemoprevention on anemia, although this is of little significance for PMC given SP’s short duration of protection [2]. Model estimates of all-cause mortality metric could capture PMC’s impact on anemia, but we did not explore this systematically. For the same reason, we did not report cost-effectiveness per disability-adjusted life-year (DALY) averted. We acknowledge that DALY estimates provide a summary of disease burden [3, 52, 53, 57], despite debates around the utility of this measure [58]. Our cost-effectiveness analysis aimed only to assess cost savings against malaria morbidity. While the study supports understanding trends in representative settings, it is not necessarily suited to inform the national or sub-national tailoring of PMC [58–60]. Finally, the current model does not estimate other, potentially positive secondary, off-target effects of SP on bacterial or fungal infections [61] that may increase the overall health benefit of PMC and PMC+.

In conclusion, we found a modest but significant public health impact and favorable cost-effectiveness for both PMC with seven SP doses and our proposed age-expanded PMC+ with nine SP doses. We identified a small risk of age-shifted malaria burden in some modelled scenarios especially for severe cases, but found that this could be mitigated with increased treatment of clinical malaria through better access to healthcare. Our results suggest that expanding the age groups for current PMC (from 24 to 36 months) could balance any reduction in impact due to partial SP-resistance, protecting the intervention’s effectiveness until new or more efficacious alternative drugs become available. Altogether, we have demonstrated that expanding age-eligibility for PMC could lead to greater effectiveness and greater cost-savings than the current deployment of PMC. Further empirical data on the feasibility and sustainability of current and age-expanded PMC deployment through either EPI or other alternative delivery channels will be essential to complement our model-based findings.

## Online Methods

### Modelling malaria transmission and interventions

#### Description of the mathematical disease model

The natural history, epidemiology, transmission dynamics of the *Plasmodium falciparum* parasite and the impact of interventions was simulated using OpenMalaria (https://github.com/SwissTPH/openmalaria/wiki), an ensemble of validated, open-access, individual-based stochastic models of malaria epidemiology [4]. It comprises of sub-model components that describe essential aspects of malaria epidemiology and interventions, including chemoprevention, case management and vector control for district, sub-district and village level population sizes. Briefly, the infection in human host is simulated as a discrete, stochastic process grounded in parasite life-stages and infection biology. The parasite density is central to describing the progression of malaria within a human-host and the effects of malaria interventions. It is linked to models of interventions and health systems. Different variants of OpenMalaria capture varying assumptions about malaria pathophysiology, transmission, the effects of comorbidity, and the effects of anti-malarial immunity acquisition and its decay. As fully described previously, all models have been fitted to field data across sub-Saharan Africa [62–64].

We applied the Molineux’s within host model variant of OpenMalaria. This model variant mechanistically depicts the time-course of asexual blood stage parasite dynamics based on the interplay between parasite growth rates and different types of host immunity assumptions following a single inoculation in an individual human host [28, 64]. This model incorporates explicit compartmental pharmacokinetic/pharmacodynamic (PK/PD) model component of drug effect in target populations [28, 64]. We specified different thresholds of the half-maximal effective concentration (EC50) to simulate varying prophylactic periods of SP for different drug sensitivity assumptions: either wild type or prevalent quadruple mutation in *Pf*dhfr and *Pf*dhps genes (dhfr-51I, dhfr-59A, dhfr-108A, and dhps-437G) [28, 39, 64]. In our model, baseline immunity was assumed in the intervention age, as we do not fully understand and could not monitor development of maternal immunity due to possible in-utero exposure to malaria or post-natal decay over time

The level of infectivity to the mosquito vector depends on the gametocyte densities in human hosts extrapolated from blood-stage parasite densities after including a lag period. The vector model component describes the plasmodium life cycle of mosquitos and its transmission probability to humans, based on a seasonally forced annual entomological inoculation rate (EIR). The model’s full characteristics, including malaria case definitions and extent and time course of intervention effects are summarized in the supplementary materials (**supplementary section 1.1 and Table S1**).

#### Description of pharmacological model of drug impact

The parasite life-stage at which SP exerts its antimalarial effect is not well characterized. Therefore, we modelled three hypothesized mode of action [42, 44] scenarios using previously validated parameters: (1) a dominant blood-stage action that clears existing infections, with a duration of action characterized by the half-maximal effective concentration based on the parasite genotype or (2) a liver-stage prophylactic action that prevents new infections as described by a Weibull decay function, or (3) combined blood-stage and liver-stage actions, assuming each effect is independent [ 38, 39] (**Table 1**). The protective efficacy against malaria morbidity under each of these assumptions was compared to empirical data ranges [1, 2, 37, 49] to identify the mode of action that best matched the reference, as briefly described below (further details provided in **supplementary section 1.2 and Tables S2 and S3).**

**Table 1.** Simulation parameters.

| Parameter | Description | Value |
| --- | --- | --- |
| Transmission pattern | Distribution of malaria incidence throughout the year, correlated with average rainfall and temperature | 1. Archetypal, constant perennial<br>2. Archetypal, exemplar sub-perennial [31] (adapted from rainfall pattern in Mozambique) |
| Transmission intensity | Entomological inoculation rate (EIR), interpreted as the number of infectious mosquito bites received yearly by a human host | 4, 8, 16, 32, 64, 128, 256, 512, 1024 |
| *Plasmodium falciparum* prevalence (*PfPR*<sub>2-10</sub>) | Test-positive patent malaria cases among 2-10 year-olds using microscopy diagnostic | *PfPR*<sub>2-10</sub> calculated as model output based on patent infection |
| Case management coverage (%) | Access to effective treatment against clinical malaria incidence, expressed as probability within 14-days post-infection (converted to a probability for a 5-day period and rounded to whole numbers) (%) | Low: 10 (5) %<br>Medium: 30 (12) %<br>High: 50 (24) %<br>Very high: 80 (53) % |
| Chemoprevention coverage (%) | Proportion of eligible children receiving IPTi or PMC or PMC+ in each dosing cycle<br>1. Full coverage, used to estimate maximum achievable impact, the maximum risk of post-intervention effects<br>2. Reduced coverage, used to simulate minimum WHO vaccination targets by 2020 in all district-level populations [17, 69] | 1. 100% (full coverage setting)<br>2. 80% (reduced coverage setting) |
| Dosing schedule | Pre-specified timing of dosing linked with the expanded program of immunization delivery schedules based on child age [12]<br>1. IPTi<br>2. PMC<br>3. PMC+ | Child age (in months)<br>1. 3, 4, 9<br>2. 3, 4, 9, 12, 15, 18, 24<br>3. 3, 4, 9, 12, 15, 18, 24, 30, 36 |
| Deterministic diagnostic | Microscopy diagnostics detecting parasite density above specified threshold | 40 parasites per $\mu\text{L}$ of blood [70] |
| **Blood-stage parasite (asexual) clearing activity [46]** |  |  |
| PK parameters for SP<sup>2</sup> (one compartment model) [28, 43] | 1. Volume of distribution (L/kg)<br>2. Absorption rate constant (per day)<br>3. Elimination rate constant (per day)<br>4. Negligible concentration (mg/L) | 1. 0.29<br>2. 12.5<br>3. 0.12<br>4. 0.001 |
| PD parameters for SP<sup>2</sup> (Michaelis-Menton kinetic<sup>3</sup>) | 1. Maximum killing rate (per day)<br>2. Slope of the effect curve<br>3. Drug concentration at which half the maximum killing rate is achieved (EC<sub>50</sub>; mg/L). Impacted by parasite genotype (mutations in key genes that drive drug-sensitivity and the duration of chemoprevention) [14] | 1. 2.3<br>2. 2.1<br>3. 2.4 for quadruple *Pf dhfr* (encoding dihydrofolate reductase)+ *Pf dhps* (encoding |
|  |  | dihydropteroate synthase) mutant genotype conferring partial SP-resistance (duration of protection ~35 days [39, 48])<sup>4</sup><br>0.5 for fully SP-sensitive wild type (duration of protection ~42 days [39, 48]) |
| **Liver-stage prophylactic activity [38]** |  |  |
| Initial efficacy | Probability of preventing individual infection at the time of administration | 100% |
| Decay profile | Deteriorating protective efficacy of a drug over time from its initial efficacy | Weibull function with shape parameter $k = 5.34$ |
| Half-life | Time (in days) until a drug's protective efficacy decays to 50% of the initial efficacy | 31 days |
| **Combined blood- and liver-stage activity** |  |  |
| As described for individual blood- and liver-stage activities, assuming no additional and no synergistic or antagonistic effect. |  |  |

<sup>1</sup>Characterized by having less than 60% rainfall occurring in the short period of increased seasonality.
<sup>2</sup>SP was treated as a single long-acting drug with previously validated parameterizations [39]<sup>2</sup>
<sup>3</sup>The EC50 was validated against the polymerase chain reaction (PCR) corrected cure rate measured in African children with uncomplicated falciparum malaria [45, 46].
<sup>4</sup>Quadruple mutant genotype: *dhfr-51I*, *dhfr-59A*, *dhfr-108A*, and *dhps-437G* mutations.

#### Validation of model estimated effect size

Our model, which estimated effect size (protective efficacy against clinical and severe malaria), was validated against a randomized controlled trial that was conducted in Mozambique [37], and the efficacy from a meta-analysis that captured wider epidemiological and clinical settings across Africa [2]. The trial site was simulated by replicating the trial’s characteristics in OpenMalaria: deployment of SP in infants, seasonality, diagnostics, and follow-up durations (**supplementary section 1.2 and Table S2**).

#### Simulation scenarios

Both PMC and PMC+ were administered as per WHO recommendations [1]: age-patterns of severe malaria in the control cohort, and potential contacts with routine EPI [6] (**Table 1, supplementary Fig. S1**). The half-maximal effective drug concentration (EC50) was used to determine the population-level prophylactic period of SP [28, 46]. Accordingly, the duration of prophylactic protection was reduced from 42 days in the fully SP-sensitive to 35 days in partially SP-resistant setting [39]. Disease progression and pathogenesis was simulated as previously described (**supplementary Table S1**). Case management was implemented by artemether-lumefantrine (AL) [65] based on its dominant use as first line treatment for clinical malaria cases [1, 2, 66].

PMC is recommended across perennial settings without any specification of additional variations in seasonal transmission [1, 29]. PMC is thereby likely to be used also in sub-perennial settings [29]. We applied uniform distribution of the entomological inoculation rate over the year to model an archetypal constant perennial transmission. Our sub-perennial seasonality patterns were adapted from rainfall patterns in parts of Mozambique, as this region has hosted or has ongoing PMC implementation and scale-up campaigns [12, 13]. Sub-perennial transmission was adjusted to distinguish it from a typical seasonal setting with less than 60% transmission in four to five consecutive months [31] (**supplementary section 1.3, Table S4 and Fig. S1a**). A two-term Fourier series expansion was used within OpenMalaria to transform these seasonal profiles to a rate of daily infectious bites (https://swisstph.github.io/openmalaria/fourier). Majority of biting was assumed to occur indoors (90%), as considered relevant for infant and young children. **Table 1** summarizes other modelling assumptions including setting characteristics, which were modelled using a full factorial design.

#### Estimation of protective efficacy

All outcomes were reported five years after PMC(+)’s rollout to capture the impact on both intervention and follow-up ages (Equation 1).

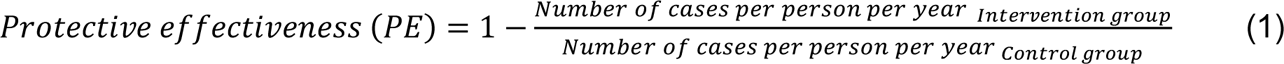

#### Estimation of post-intervention effects

We examined any possible post-intervention effects: the incidence of delayed malaria, and cumulative malaria cases (net or total program impact) over the intervention and follow-up ages, as per WHO’s recommendation [19]. To explore the maximum risk of potential age-shifting these indicators were monitored under full (100% in all cycles) PMC(+) coverage across settings.

#### Cost-effectiveness analysis

The cost of PMC or PMC+ implementation [1] was calculated based on the cost per SP dose delivered for a complete course for a hypothetical intention-to-treat cohort of 1,000 children under 36 months. The cost per dose delivered was informed by comprehensive meta-analysis that covered clinical trials and implementation studies and reflected economic and health system scenarios from across Africa (**supplementary Table S5**) [49]. The mean cost was inflation-adjusted using a US dollar inflation rate calculator (http://www.usinflationcalculator.com), leading to a mean inflation-adjusted cost of US$0.29 per dose delivered in 2024 (**supplementary section 1.4, Table S5**). A discounting of 3% on cost and no discounting on outcomes was applied, following WHO’s Choosing Interventions that are Cost-Effective (WHO-CHOICE) 2021 update [60]. We used clinical-cases-averted as the measure of effectiveness, in line with some other studies [53, 59].

The incremental cost-effectiveness ratio (ICER) against clinical or severe malaria was calculated for implementing either PMC or PMC+ relative to a counterfactual scenario of no chemoprevention (only case management) (Equation 2).

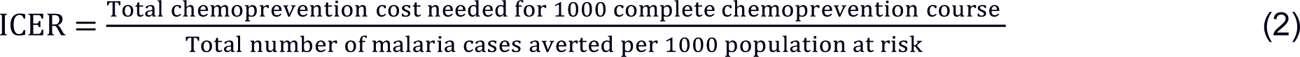

Similarly, incremental cost-effectiveness of PMC+ was assessed compared to that for PMC (ICER_PMC+/PMC_) (Equation 3).

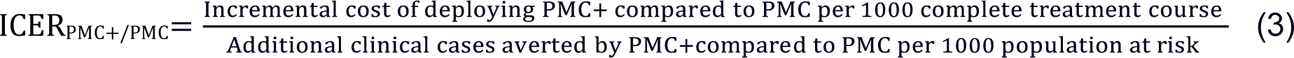

As a secondary analysis, we calculated cost-effectiveness trends that included cost savings. The costs of case management were estimated by giving a maximum of 18 (six doses with three tablets in each) artemether-lumefantrine (AL) for managing clinical malaria in children having body weight 25 to 35 kg [67]. The cost of AL was extracted from a study for Mozambique, assuming a cost of US$0.38 (Management Sciences for Health price in 2003) for a pack of 16 tablets or capsules [68] and applying an inflation-adjustment to yield a 2024 value of US$0.65. The total cost was calculated by subtracting the cost of treating one clinical case from the cost of chemoprevention to avert one case (Equation 4).

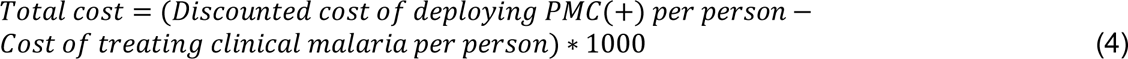

## Data and code availability

The source code for the applied individual-based model OpenMalaria is publicly available at https://github.com/SwissTPH/openmalaria (https://doi.org/10.5281/zenodo.10534022) and the detailed documentation at https://github.com/SwissTPH/openmalaria/wiki. The archived version of the model simulation and data analysis codes are available at https://doi.org/10.5281/zenodo.12721515. The R scripts and source data used for production of figures presented in this paper can be found at https://doi.org/10.5281/zenodo.12722070.

## Acknowledgements

This study was funded by the Bill & Melinda Gates Foundation (INV-025569 to MAP). LBM and SK acknowledge support from Bill & Melinda Gates Foundation INV-002562 to MAP. TM and MAP acknowledge support from the Swiss National Science Foundation (SNF Professorship PP00P3_170702 and PP00P3_203450 to MAP). JJM was employed at Medicines for Malaria Venture (MMV) during study period. We acknowledge all support and advice from all members of the Disease Modelling Research unit of the Swiss Tropical and Public Health Institute. We acknowledge technical support from Dr. Andrew J Shattock. We thank Daria Hofer for the project management support. All analysis and calculations were performed at sciCORE (https://ood.scicore.unibas.ch/) scientific computing centre at University of Basel.

## Author information

**Swiss Tropical and Public Health Institute, Allschwil, Switzerland**

Swapnoleena Sen, Lydia Braunack-Mayer, Sherrie L Kelly, Thiery Masserey, and Melissa A Penny

**University of Basel, Basel, Switzerland**

Swapnoleena Sen, Lydia Braunack-Mayer, Thiery Masserey, Joerg J Moehrle and Melissa A Penny

**Medicines for Malaria Venture, Geneva, Switzerland**

Joerg J Moehrle

**Telethon Kids Institute, Perth, WA, Australia**

Josephine Malinga and Melissa A Penny

**Centre for Child Health Research, The University of Western Australia, Perth, WA, Australia**

Josephine Malinga and Melissa A Penny

## Contributions

SS and MAP designed the study. SS developed and performed analyses. SLK supported the cost-effectiveness analysis. LBM, SLK, TM, JM, JJM and MAP validated the model, analyses, and results. SS drafted the manuscript and prepared figures. All authors contributed to interpreting the results and making edits to the draft and final manuscript and gave their final approval for publication.

## Ethics declarations

Joerg J Moehrle was employed by Medicines for Malaria Venture. Melissa A. Penny was a member of the WHO Guidelines Development Group for Malaria Chemoprevention in 2020-2021. All other authors declare no competing interests.

## Supplementary Information

### 1. Methods

#### 1.1 Model of malaria transmission and control (OpenMalaria)

We assessed the potential public health impact of the updated perennial malaria chemoprevention (PMC) guidelines [1, 2], and of a proposed age-expanded PMC program. We did this by applying OpenMalaria, an open-source, validated individual based model of malaria epidemiology and control [3]. In this model, the asexual blood stage parasite density in human determines the progression of the disease, and it can be reduced by naturally acquired immunity (either variant-specific or variant-transcending or adaptive) in the absence of interventions. The extent and time course of an intervention’s effect depends on its mechanism of action (**Table S1**). The incidence of malaria clinical cases (symptomatic, but uncomplicated), severe cases (following WHO definition [4, 5]), malaria related deaths and all-cause mortality are tracked for each timestep over the pre-intervention (baseline), intervention and follow-up time periods.

The vector model within OpenMalaria describes the *Plasmodium* life cycle in mosquitos and the probability of transmission to a human host based on their number of infectious mosquito bites, tracked in five day timesteps. The annual entomological inoculation rate (EIR) is seasonally forced by applying a Fourier transformation (https://swisstph.github.io/openmalaria/fourier), and is dependent on the infection sub-model used. We did not focus on the spatiotemporal heterogeneity of the vector species, including their host attractiveness pattern by age or their changing dynamics [6]. Instead we modeled a generic vectoral pattern in this work by simulating the entomological characteristics of *Anopheles gambiae,* one of the most widespread species across Africa [6, 7].

To address multiple research question covering a range of public health applications, 14 different variants of OpenMalaria have evolved. Each model variant captures varying assumptions about malaria pathophysiology, anti-malaria immunity acquisition and its decay, the characterization of intervention effects, patterns of *Plasmodium* parasite transmission, and effects of comorbidity, among others [8]. We applied Molineux’s within-host model variant in this study. This particular model variant mechanistically describe the time course of infection progression from asymptomatic to clinical malaria and to either severe outcome(s) or recovery [9]. It incorporates explicit population pharmacokinetic and pharmacodynamic (PK/PD) sub-model components to depict the time course of a drug’s (either chemoprevention or treatment) exposure-response relationship in specific target population following a clinically relevant dosage. The key model components and characteristics used in this study are detailed in **Table S1**; The full model description is recorded at https://github.com/SwissTPH/openmalaria/wiki.

**Table S1.** Overview of OpenMalaria model, adapted from previous publications [10, 11].

| Key modeled processes | Description and underlying assumptions | References |
| --- | --- | --- |
| **Malaria disease biology and epidemiology** |  |  |
| Infection in individual human host | Exposure is extrapolated from number of infectious mosquito bites over the year, specified by the entomological inoculation rate (EIR)<br>Seasonality pattern characterized by the monthly distribution of EIR<br>Correlates to availability based on age-dependent body surface area | [3, 12] |
| Progression of infection determined by asexual parasite density and acquired anti-malarial immunity | Explicit mechanistic within-host model of asexual parasitaemia<br>Various natural immunity (innate, variant-specific, variant-transcending, adaptive can reduce asexual parasite density in human) acquired over multiple age-dependent infection history<br>Both pre-erythrocytic liver-stage and blood-stage immunity decays exponentially over time<br>Duration of infection follows a log-normal distribution obtained by fitting to malaria therapy dataset in the original model (collated by Marsh and Snow [13]) | [3, 9, 12, 14-16] |
| Clinical cases, morbidity, mortality and anaemia | Clinical malaria episode depends on human host parasite density and their pyrogenic threshold, which evolves over time depending on the individual exposure history<br>Patent malaria detected by threshold based on the type of diagnostic used (for instance, 40 parasites/μL of blood by microscopy detection was specified in this study)<br>Infections can be uncomplicated clinical cases, or can evolve to severe cases, based on parasite density and the pyrogenic threshold in each individual host<br>Severe disease can also be induced by typical age-based comorbidities<br>A proportion of the severe cases leads to deaths, resulting in direct malaria-related mortality<br>Indirect mortality classified as deaths in patients not diagnosed as malaria deaths but would not occur without a malaria exposure<br>All-cause mortality captures deaths resulting from both direct, indirect malaria-related causes, as well as hospitalization deaths, and thereby captures also impact of comorbidity (such as anemia) | [3, 17-19] |
| **Transmission setting characteristics** |  |  |
| Population age structure | Flexible and informed by health and demographic surveillance data from Tanzania | [19, 20] |
| Transmission seasonality | Specified by monthly EIR distribution<br>In the absence of interventions seasonality is reproduced each year | [12, 21] |
| Transmission from infected humans to mosquitoes | The level of infectivity depends on the density of the sexual form of parasites (gametocyte densities) present in human hosts extrapolated from blood-stage parasite densities, including a lag period<br>Mosquitoes become infectious follows a binomial distribution | [3, 22, 23] |
| Entomological setting | Mosquito lifecycle and behavior towards human and non-human hosts (such as biting and resting) is embedded in a dynamic entomological model capturing the cycle of mosquito oviposition<br>Multiple vector species can be simulated simultaneously<br>Each variant-specific parasite has its own multiplication rate drawn from a Normal distribution with mean of 16 | [24] |
| **Intervention deployment characteristics** |  |  |
| Drugs and other therapeutic modalities | Interventions (small molecule drugs, vaccines, monoclonal antibodies, etc.) can act at different parasite life cycle stages | [16, 25] |
|  | (either by impacting on survival and emergence of asexual liver-stage or killing blood-stage parasites, blocking transmission) |  |
| Case management | Modelled through a decision tree-based approach, which determines treatment implications depending on the occurrence of clinical cases<br>Its representation includes specification of diagnostic tests, effects of treatment, case fatality, case sequelae, and cure rates, influenced by the diagnostic threshold and access to care | [26] |
| Intervention deployment characteristics (as applied to model chemoprevention,) | In addition to treatment of clinical malaria episodes, chemoprevention interventions were deployed in the model either:<br>Continuously: to individuals at pre-specified ages over a specified time, and with specified coverage level (such as, administering age-targeted perennial malaria chemoprevention (PMC) administration modelled in this study)<br>Timed: at specified time points (annually or over multiple years) to targeted groups over several cycles and at specified coverage levels, typically seasonally-targeted (such as, seasonal malaria chemoprevention (SMC) delivery)<br>Interventions can be deployed by enrolling individuals into cohorts and tracking cohort outcomes, facilitating clinical trial simulation | [16, 27-29] |
| Vector control | Probability of infection in human host can be reduced by vector control interventions<br>Interventions include long-lasting insecticide-treated nets (ITN), indoor residual spraying (IRS), house screening, baited traps, mosquito repellents, and push-pull | [24] |
| **Simulation characteristics and model variants** |  |  |
| Timestep | Simulations are tracked for every 5 day timestep<br>Tracked output metrics (for instance, clinical cases) from each simulated scenario can be extracted per timestep (5 days), monthly, quarterly or yearly values | [19] |
| Model variants | Varying assumptions for immunity decay, treatment effect, and heterogeneity in transmission are captured in 14 model variants<br>In this study Molineux within-host model adapted including explicit PK/PD characteristic of the intervention drugs for chemoprevention drug sulphadoxine-pyrimethamine (SP), and treatment drug artemether-lumefantrine (AL) | [9, 16] |

#### 1.2 In-silico trial: validation of model estimated effect size

An in-silico trial was conducted to ensure that our model captures SP’s age-targeted prophylactic impact over a range of epidemiological and clinical scenarios (**Table S2** [1, 30]), and to identify SP’s parasite life-stage specific mode of action [30, 31]. We simulated the trial site settings, such as seasonality in Manhiça (increased rainfall from November to April) and malaria case definitions [31, 32]) (**Table S2**, **Fig. S1**). Microscopy diagnostics were applied to detect parasite densities of 40 parasites per µL of blood [33]. The protective efficacy (PE) against clinical and severe malaria was estimated in infants (0-12 months of age) based on incidence rate ratio [34, 35] 12 months following the intervention’s start (Equation. 1). This allowed initial trial participants to complete a full therapeutic course. Since our model monitors the entire population at risk rather than tracking individual outcomes, we assessed the PE based on relative reduction of the incidence rate ratio instead of using the Kaplan-Meier method. Our method was consistent with the reference meta-analysis (**Table S3**) [30]. Also, it was not necessary to adjust for any confounding or sampling errors since the entire population was monitored.

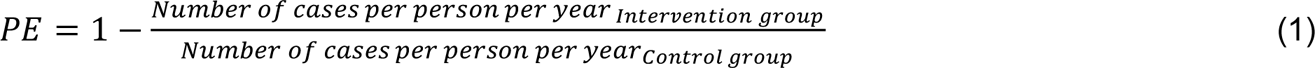

**Table S2.** Overview of the randomized controlled trial and in-silico study design [31].

| Parameter | Randomized controlled trial | In-silico trial using OpenMalaria |
| --- | --- | --- |
| Trial design |  |  |
| Timeline | September 2002 to February 2004 |  |
| Location | Manhiça, Mozambique |  |
| Participant characteristics | Number of participants: 1,503<br>Inclusion criteria: infants (aged 3 months at first dose); permanent residence | From a village population size of 10,000 mean number of 250 infants (aged 3 months at first dose) |
| Interventions | Sulphadoxine-pyrimethamine (SP) was given at 3, 4, and 9 months of age; administered according to total body weight: <5 kg, 1/4 tablet; 5–10 kg, 1/2 tablet; >10 kg, 1 tablet |  |
| Length of follow-up | 12 months (until 24 months of child age) |  |
| Setting characteristics |  |  |
| Transmission intensity | Entomological inoculation rate (EIR) of 38 infective bites per person per year | Mean EIR of 38 infective bites/person/year [35-40]; 90% outdoor biting rate |
| Seasonality | Moderate perennial setting with a rainy season from November to April |  |
| Case definitions | Clinical malaria- axillary temperature of $\geq 37.5^{\circ}\text{C}$, together with asexual *P. falciparum* parasitaemia of any density.<br>Severe malaria-WHO definition [4, 5] | Clinical malaria: axillary temperature of $\geq 37.5^{\circ}\text{C}$, together with asexual *P. falciparum* parasitaemia $\geq 40/\text{uL}$ of blood (microscopy); after 6 timesteps (30 days) a person could get re-infected<br>Severe malaria-WHO definition [4, 5] |
| Case management | Uncomplicated malaria was treated with 7 days of oral quinine, if IPTi had been administered within the preceding 2 weeks<br>During the course of the study, first line treatment for uncomplicated malaria changed from chloroquine to combination therapy with Amodiaquine and SP | Applied simple blood-stage parasite clearance model with 100% efficacy (at each 5 day timestep) |
| Analysis |  |  |
| Method | Primary efficacy analysis conducted in the Intent-to-treat (ITT) cohort<br>The protective effect (PE) was estimated from the hazard ratio (HR) as $\text{PE} = 100 * (1 - \text{HR}) \%$<br>Secondary analyses included assessments of multiple episodes of malaria using Poisson regression models | Primary efficacy analysis conducted in the ITT cohort<br>The protective effect (PE) was estimated from incidence rate ratio after complete intervention cycle $\text{PE} = 100 * (1 - \text{IRR}) \%$ at stable post-intervention time point |
| Result | Primary results by ITT, secondary analysis per-protocol (PP) showed comparable results<br>All episodes of clinical malaria were reduced by **22.6% (1.6 – 39.2%)**<br>All episodes of severe malaria were reduced by **12.7% (-17.3% – 35.1%)** | Primary results by ITT, secondary analysis per-protocol (PP) showed comparable results<br>All episodes of clinical malaria were reduced by **22.7% (16.2% – 29.1%)<sup>2</sup>**<br>All episodes of severe malaria were reduced by **18.1% (10.7% – 25.5%)<sup>2</sup>** |
| <sup>1</sup>Results indicate the median (interquartile range) of protective efficacy from modeling SP's blood-stage specific mode of action in OpenMalaria [36] |  |  |

**Table S3.** Meta-data ranges that informed WHO’s PMC guideline update [30, 37].

| Outcome metric | Effect size (incidence rate ratio) | Certainty level of evidence |
| --- | --- | --- |
| Clinical malaria | 0.78 [0.69–0.88] | Moderate |
| Severe malaria | 0.92 [0.47–1.81] | Low |

Additionally, we compared efficacy following intention-to-treat (ITT) and per-protocol (PP) dosing schedules as per the trial (**extended data Fig. E6**). ITT included all participants who received any number of SP doses, while the PP analysis was limited to participants who received all age-specified doses. The latter is similar to vaccine schedules, where a follow-up booster dose is usually given only if all prior doses were administered.

#### 1.3 Scenario design

All simulations were run using ten random seeds to capture stochastic variation. Trends and dispersions for all outcome matrices were summarized by calculating the median and interquartile range respectively to account for outliers from a non-normal distribution [38].

##### Seasonality characteristics

We modeled two archetypal transmission settings: (1) a constant perennial transmission, and (2) an exemplar sub-perennial (such as observed in the replicated trial site) [39, 40]). For the perennial transmission we applied uniform distribution of the entomological inoculation rate over the year. Seasonality values for the sub-perennial transmission was extracted from the distribution of average rainfall. Since Mozambique’s original rainfall across data Mozambique exhibited a rather strong seasonal transmission (over 60% of total annual rainfall in four consecutive months [40], **Fig. S1**), we adapted the distribution of transmission intensity by adjusting the Fourier transformation coefficients within OpenMalaria (https://swisstph.github.io/openmalaria/fourier, **Table S4 and Fig. S1**). Thereby, the modeled scenario demarcated from the seasonal transmission profiles that are covered by seasonal malaria chemoprevention (SMC) [1]. To note, the WHO malaria terminology glossary recently incorporated an explicit category for sub-perennial seasonality (2021) [39]. Studies prior to this usually described all settings with year-round transmission as perennial [31]. For instance, in the replicated trial site, the seasonality was described as “subtropical, with a warm rainy season from November to April and a cool dry season during the rest of the year” [31], closely aligned with what is now classified as sub-perennial [39]. Overall, our modelled scenarios covered wide range of the settings where PMC is currently recommended [1, 41]

**Table S4.** Overview of model parameters for adapting archetypal sub-perennial transmission.

| Seasonality | Entomological inoculation rate rotate angle | Coefficient a1 | Coefficient a2 | Coefficient b1 | Coefficient b2 |
| --- | --- | --- | --- | --- | --- |
| Mozambique (normalized rainfall) | 0 | 1.0414 | 0.1916 | 1.2188 | 0.1078 |
| Sub-perennial (adjusted values as modelled) |  | 0.6414 |  | 0.5188 |  |

**Fig. S1.**
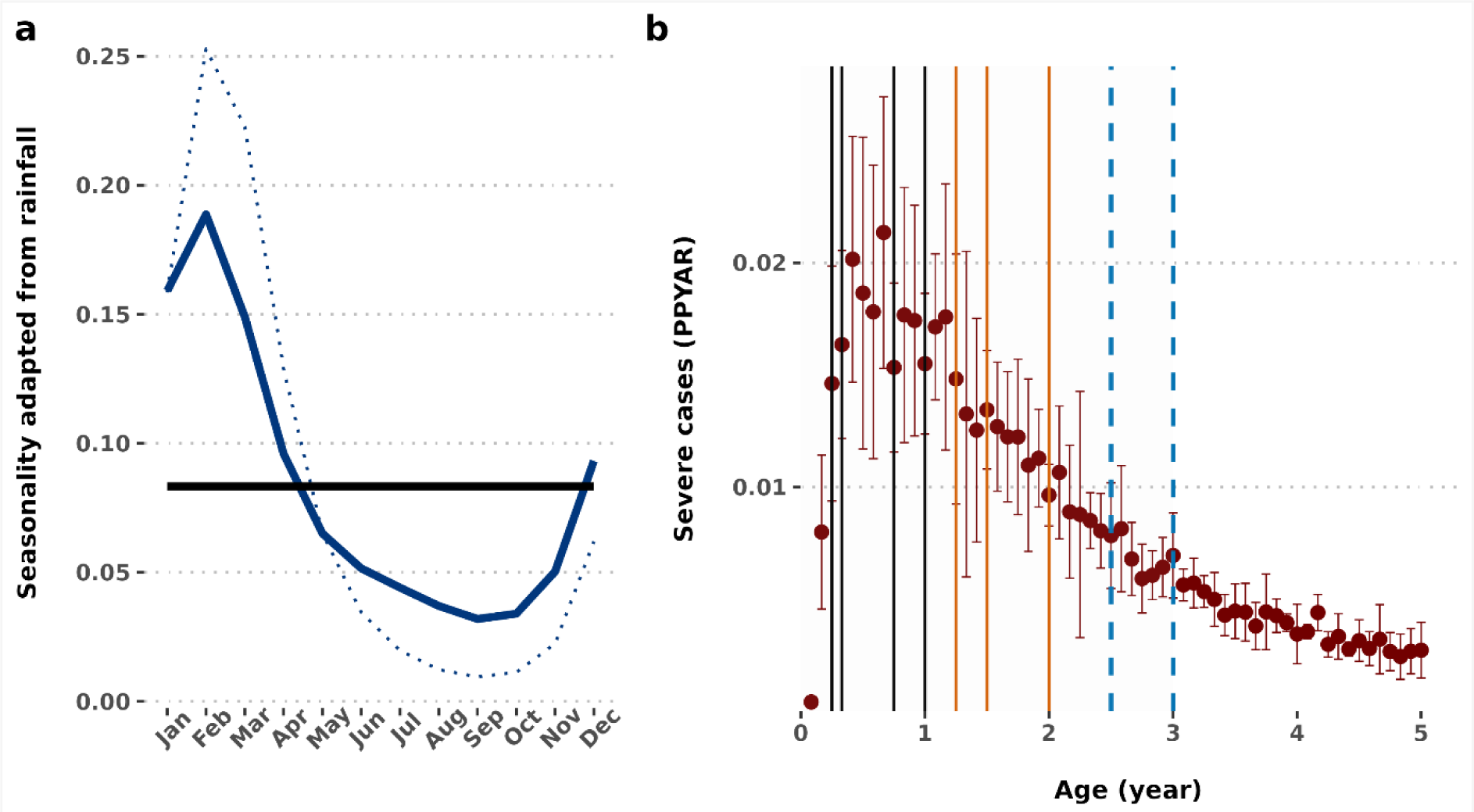
Overview of the model scenarios. The left plot in panel (a) shows seasonality patterns in archetypal perennial and sub-perennial settings. The solid black line denotes a constant perennial transmission, solid blue line shows seasonality pattern based on average monthly rainfall in Mozambique and the dotted blue line indicates the modeled exemplar sub-perennial malaria transmission as adjusted from the Mozambique rainfall pattern. **The right plot in panel (b) describes dosing schedule shaped by age-pattern of severe malaria in the control cohort, and potential contacts with routine expanded program of immunization delivery.** The red dots and error bars depict median and interquartile range of severe malaria cases. Four PMC or PMC+ doses in the first year of life (at age 03, 04, 09 and 12 months) is depicted by the solid black line, three PMC or PMC+ doses in the second year of life (at age 15, 18 and 18 months) is depicted by the solid orange lines and two additional PMC+ doses in the third year of life (at age 30 and 36 months) are depicted by the light blue dashed lines. PMC: Perennial malaria chemoprevention; PMC+: Age-expanded perennial malaria chemoprevention; PPYAR: Per person per year at risk.

#### 1.4 Cost-effectiveness analysis

The cost of PMC or PMC+ delivery using SP through the existing expanded program of immunization was extracted from a recent systematic review and meta-analysis results [37]. This incorporated estimates from controlled trials and implementation studies from across malaria endemic regions of Africa. We applied the cost of delivering SP based on the mean price inflated at their 2024 values using http://www.usinflationcalculator.com (**Table S5**) [1, 2].

**Table S5.** Costs per sulphadoxine-pyrithiamine dose delivered through the expanded program of immunization, adapted from the summary of intermittent preventive treatment in infants (IPTi) contextual factors^1^.

| Study setting (study year) | Cost per dose delivered in the study year (US\$) | Inflated cost per dose delivered in 2024 (US\$) | Reference |
| --- | --- | --- | --- |
| RCT (large scale community randomized trial): Tanzania (2005) | 0.23 (78% financial expenditure and 22% opportunity cost) | 0.37 | [42] |
| RCT (large scale pilot studies): Tanzania (2006), Mozambique (2006) | 0.13<br>0.15 | 0.20<br>0.23 | [43] |
| **Meta-analysis of multiple trial site data:** Tanzania, Mozambique, Gabon, Kenya, and Ghana (2006) | 0.13 [0.09–0.17] | 0.20 [0.14–0.26]<sup>2</sup> | [44] |
| **Implementation study:** Ghana (2007) | 0.29 (0.87 for 3 routine doses) | 0.44 | [45] |
| **Mean delivery cost** | **0.19** | **0.29** |  |

<sup>1</sup>Systematic reviews, background papers and other unpublished evidence considered in the development of recommendations, Section 4.2.2 Perennial malaria chemoprevention (PMC), WHO Guidelines for malaria, 3 June 2022 [1, 37]
<sup>2</sup>Secondary analysis with 95% confidence interval [44], built on the cost-effectiveness model [43]

## Extended data figures

**Fig. E1.**
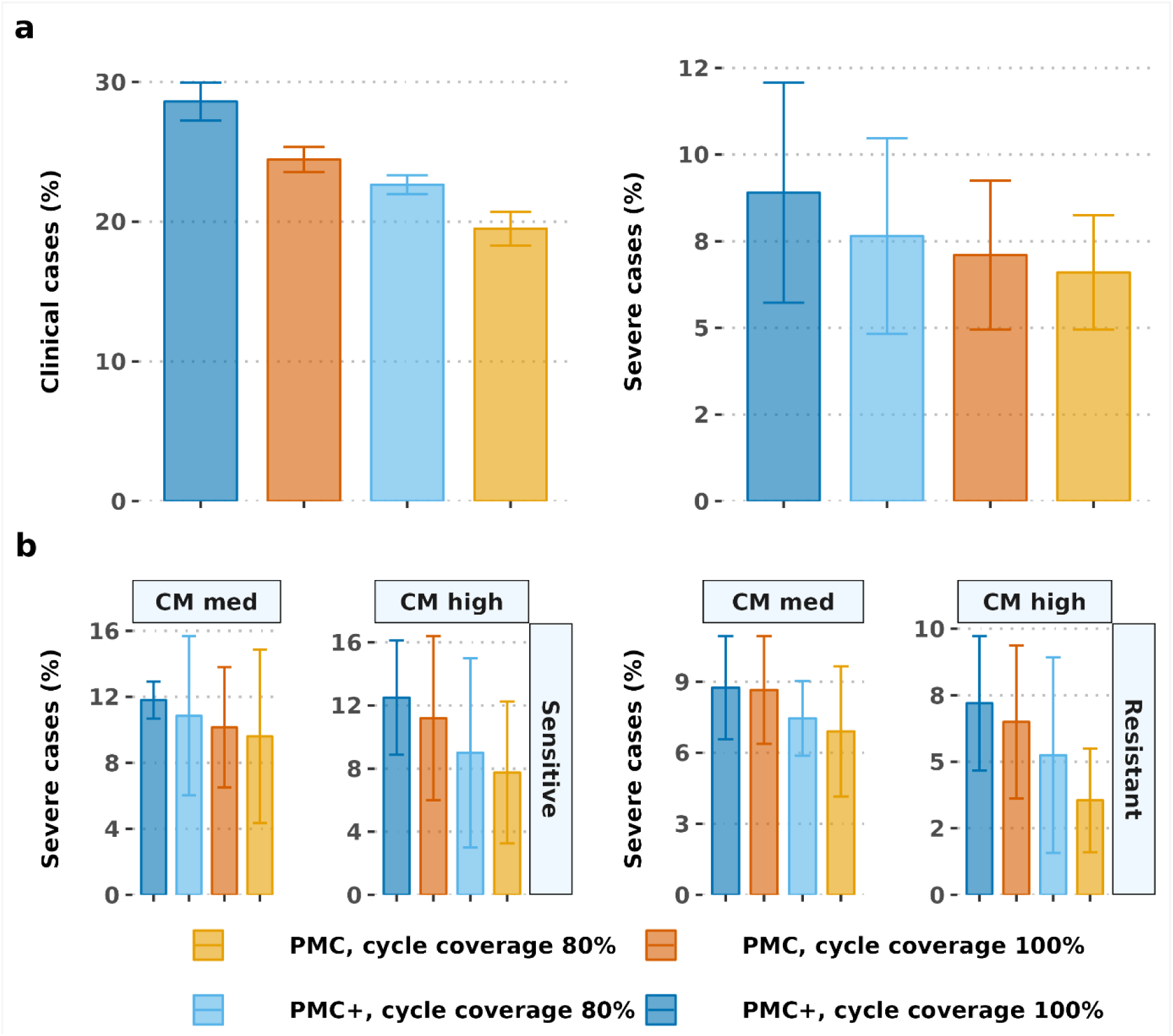
Protective efficacy against all episodes of clinical and severe malaria in the first three years of life under full (100% in each dosing cycles) vs. reduced chemoprevention coverage (80% in each dosing cycles). The top panel (a) shows the differential impact of coverage on clinical vs. severe cases. The trend for effectiveness against clinical cases were consistent across setting. Exemplar result is shown in low case management (10% probability of accessing case management within 14-days post-diagnosis) and partial SP-resistant setting. **The bottom panel (b) depicts differential trend of effectiveness against severe cases based on different case management levels and drug sensitivity assumptions.** Results are shown in settings with high baseline prevalence (*Pf*PR_2-10_ 30-59%). CM: Case management; PMC: perennial malaria chemoprevention; PMC+: Proposed age-expanded perennial malaria chemoprevention; *Pf*PR_2-10_ : P. falciparum prevalence in 2-10 year olds.

**Fig. E2.**
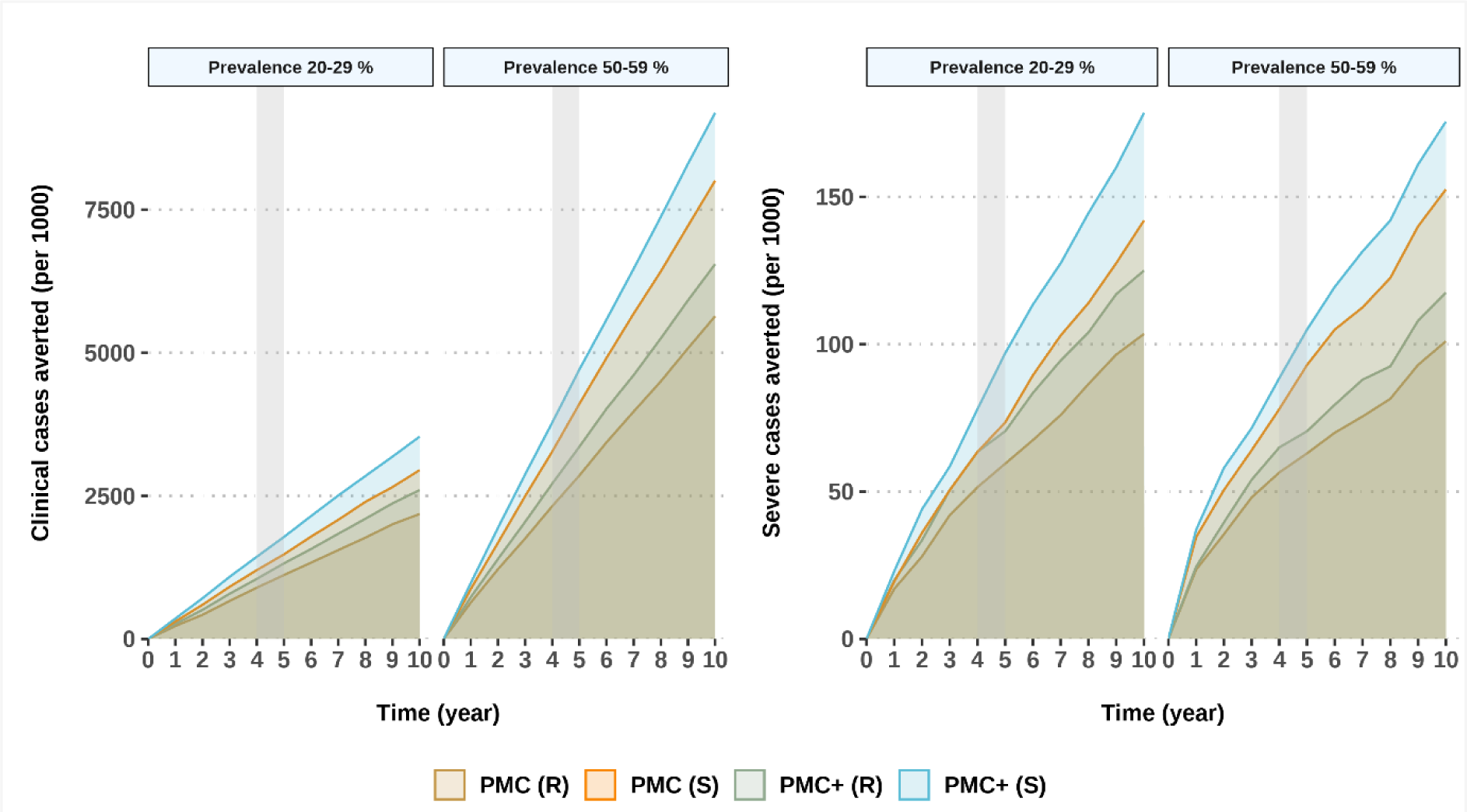
Cumulative incidence of clinical and severe cases averted by PMC or PMC+ in children under three years, over time. Results depict perennial settings with medium (P*f*PR_2-10_ 20-29%, EIR 8) to high transmission intensity (P*f*PR_2-10_ 50-59%, EIR 256), 30% probability of accessing case management within 14-days post-diagnosis, and full coverage. The grey shaded area denotes the analysis time point after PMC(+) rollout. EIR: Entomological inoculation rate; PMC: perennial malaria chemoprevention; PMC+: Age-expanded perennial malaria chemoprevention; *Pf*PR_2-10_ : P. falciparum prevalence in 2-10 year olds; R: Partially SP-resistant; S: SP-sensitive.

**Fig. E3.**
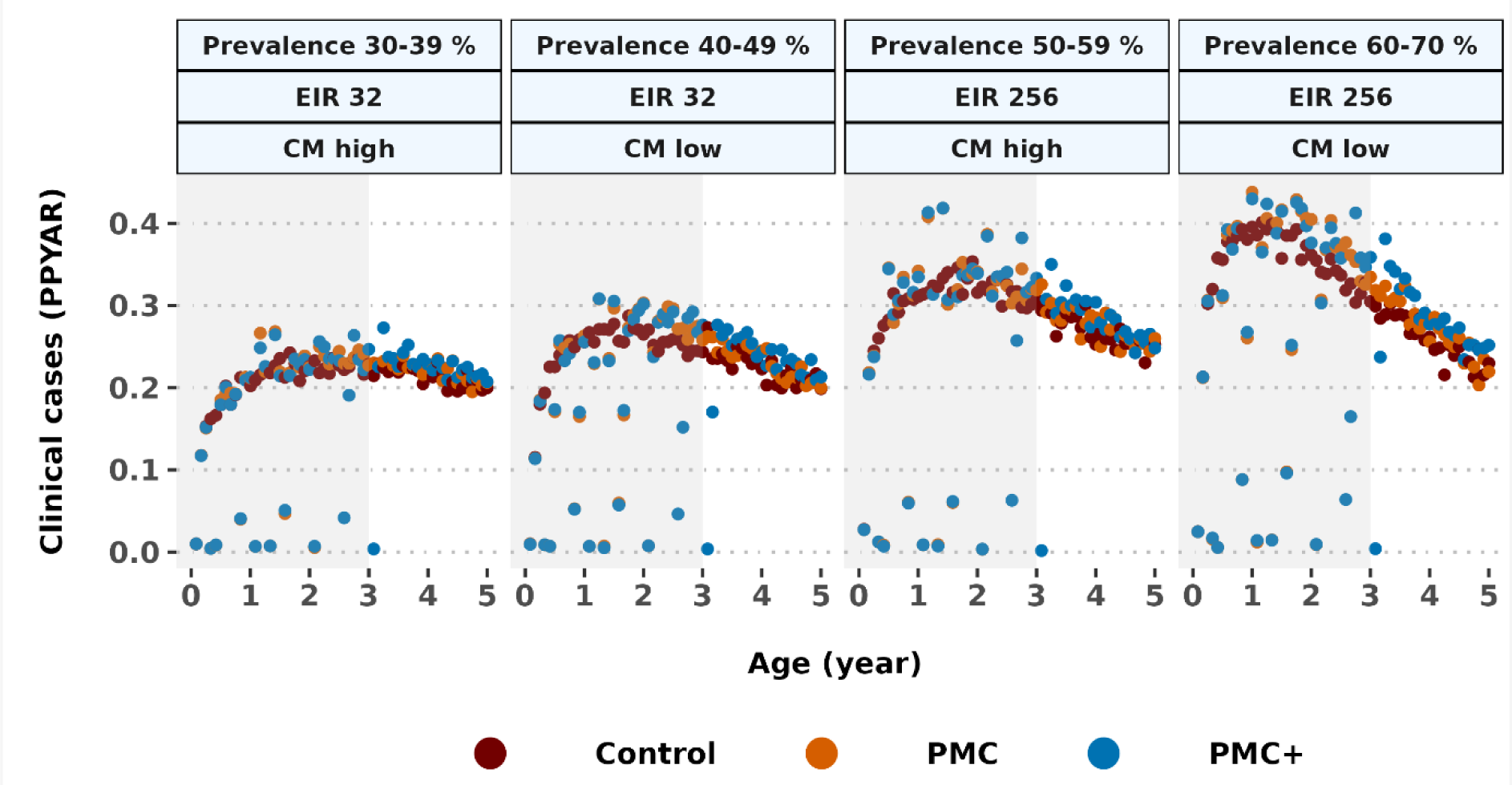
Distribution of clinical malaria incidence per age. The grey shaded area denotes intervention age. The strength of healthcare system is depicted by low (10%) and high (50%) probability of accessing case management within 14-days post-diagnosis, in partially SP-resistant (R) setting. EIR: Entomological inoculation rate; CM: Case management; PMC: perennial malaria chemoprevention; PMC+: Age-expanded perennial malaria chemoprevention; PPYAR: Per-person per year at risk.

**Fig. E4.**
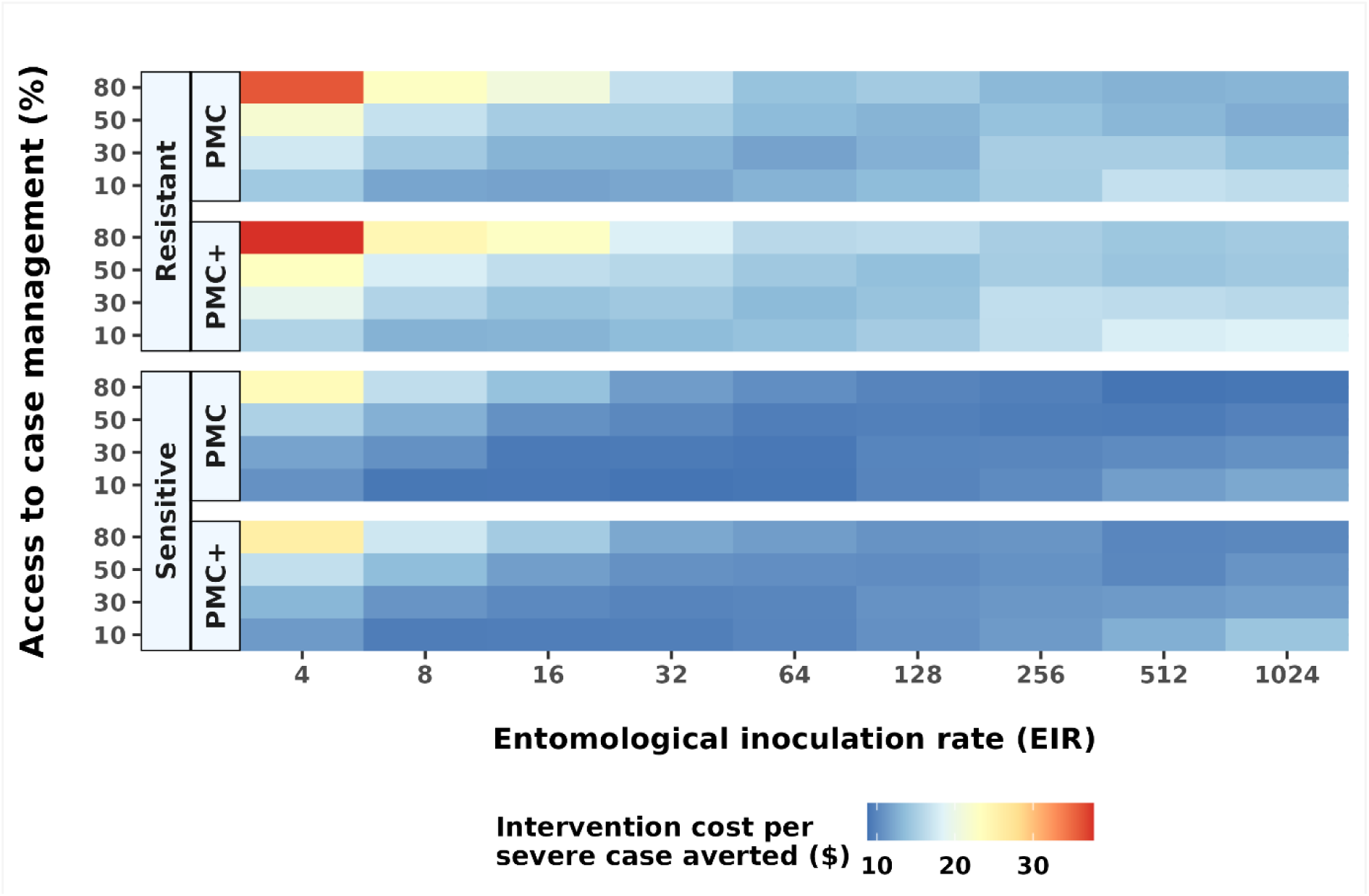
Incremental cost effectiveness against severe malaria for PMC or PMC+ in partially SP-resistant and fully SP-sensitive settings with different levels of access to treatment, compared to a counterfactual of no chemoprevention. CM: Case management; PMC: perennial malaria chemoprevention; PMC+: Proposed age-expanded perennial malaria chemoprevention; SP: sulphadoxine-pyrimethamine.

**Fig. E5.**
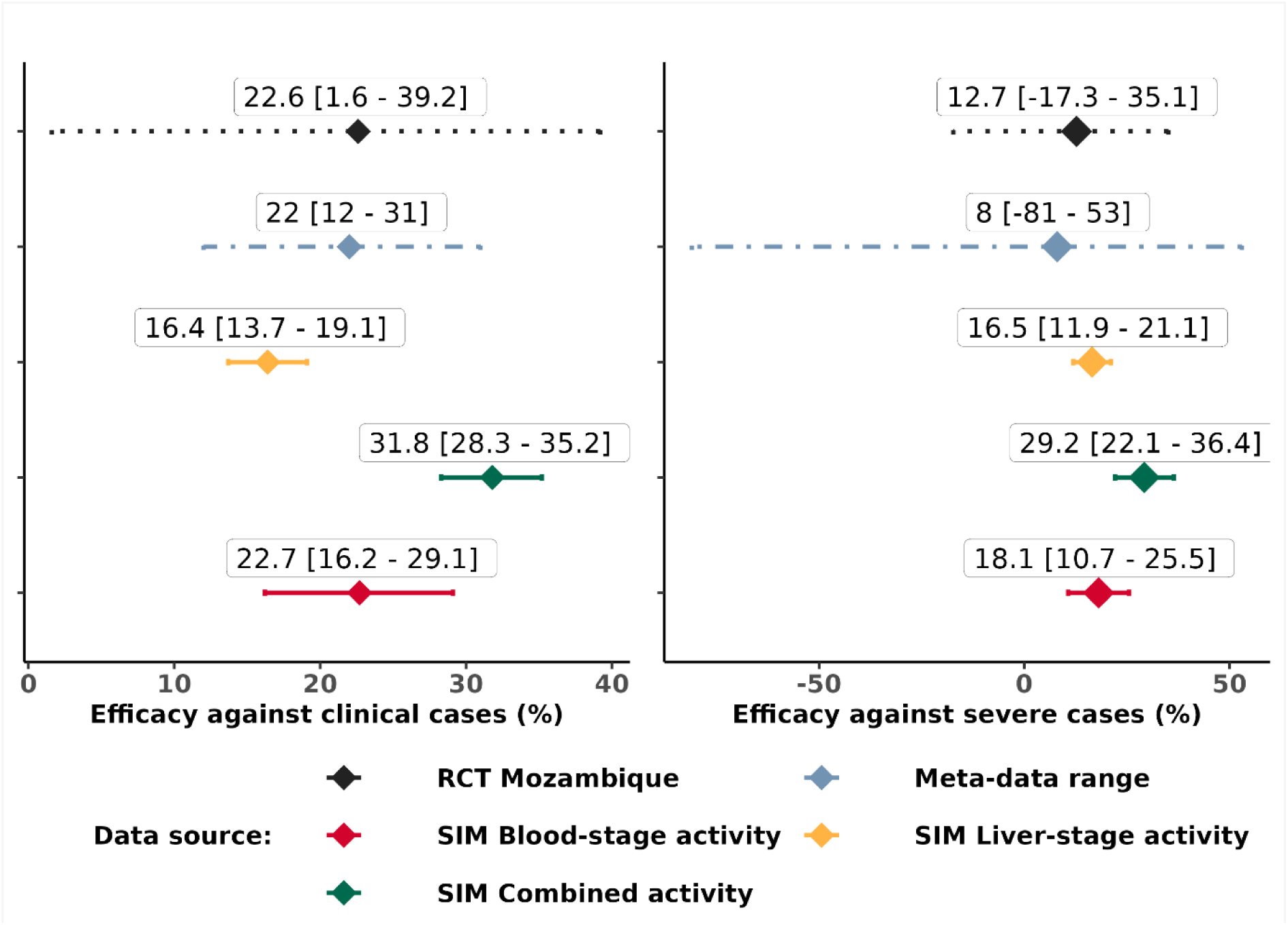
Comparison of model estimated effect size to empirical data ranges from the replicated randomized control trial (RCT), and meta-data ranges. The effect represents relative incidence rate reduction of clinical and severe malaria cases. The results are shown for three simulated (SIM) mechanisms of sulphadoxine-pyrimethamine drug action: only blood-stage activity (red lines), only liver-stage activity (yellow lines), and combined blood- and liver-stage activity (green lines). The model estimated efficacies are shown with solid lines, randomized controlled trial results with black dotted lines, and the meta-analysis results are shown with blue-grey dot-dashed lines. The results depict median and interquartile range across simulated seasonality settings having medium access to healthcare (30% probability of accessing case management within 14-days post-diagnosis), and 100% IPTi coverage at each dosing cycles. IPTi: Intermittent preventive treatment in infants.

**Fig. E6.**
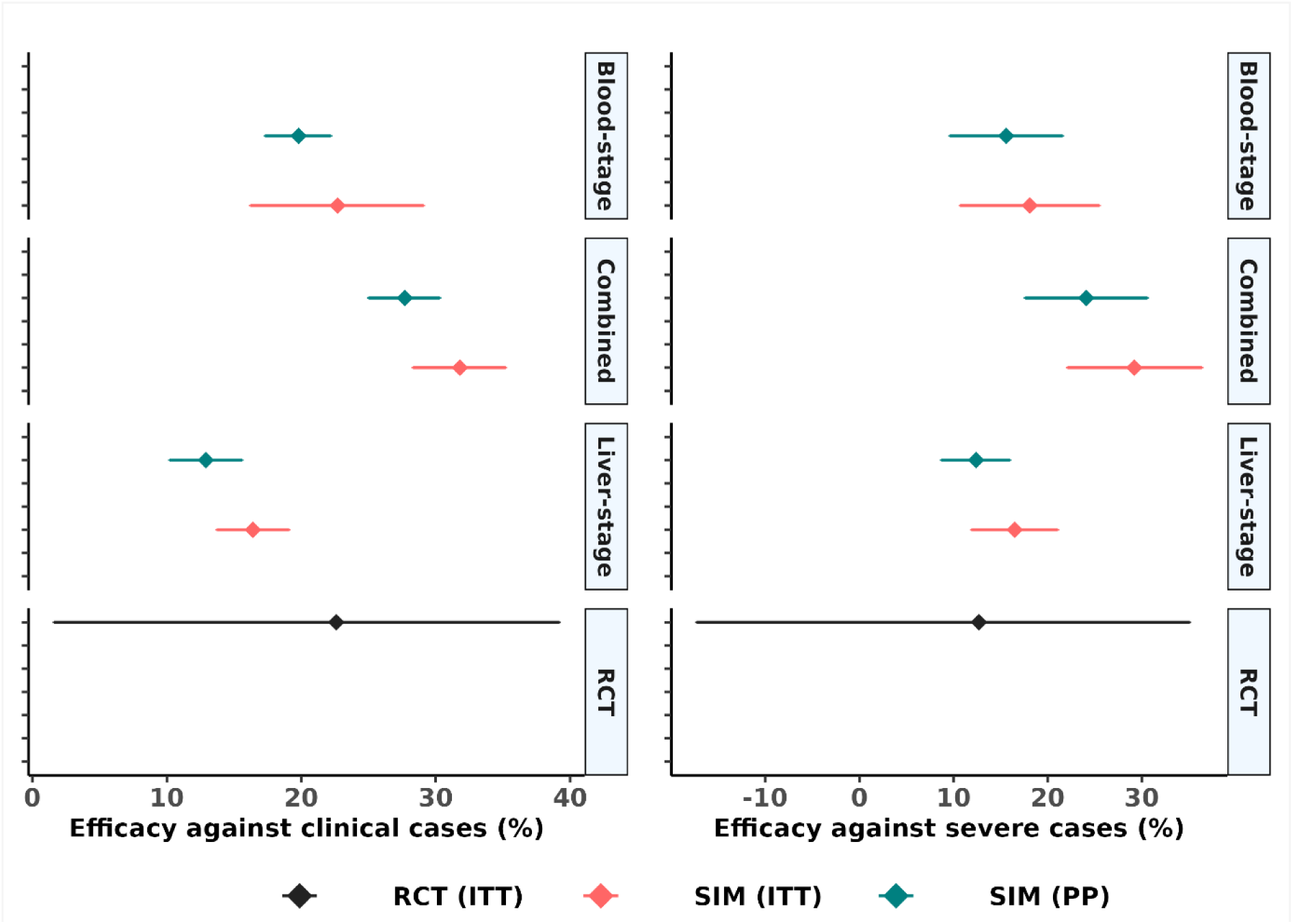
The range of protective efficacy following intention-to-treat (ITT) vs. per-protocol (PP) dosing schedule for replicated IPTi trial with sulphadoxine-pyrimethamine. Sub-panels show results from model simulated (SIM) cohort having different assumptions of parasite life-stage specific drug action, and results from the randomized controlled trial (RCT). The results depict median and interquartile range across simulated seasonality settings having medium access to care (30% probability of accessing case management within 14-days post-diagnosis), and 100% IPTi coverage at each dosing cycle. IPTi: Intermittent preventive treatment in infants.

**Fig. E7.**
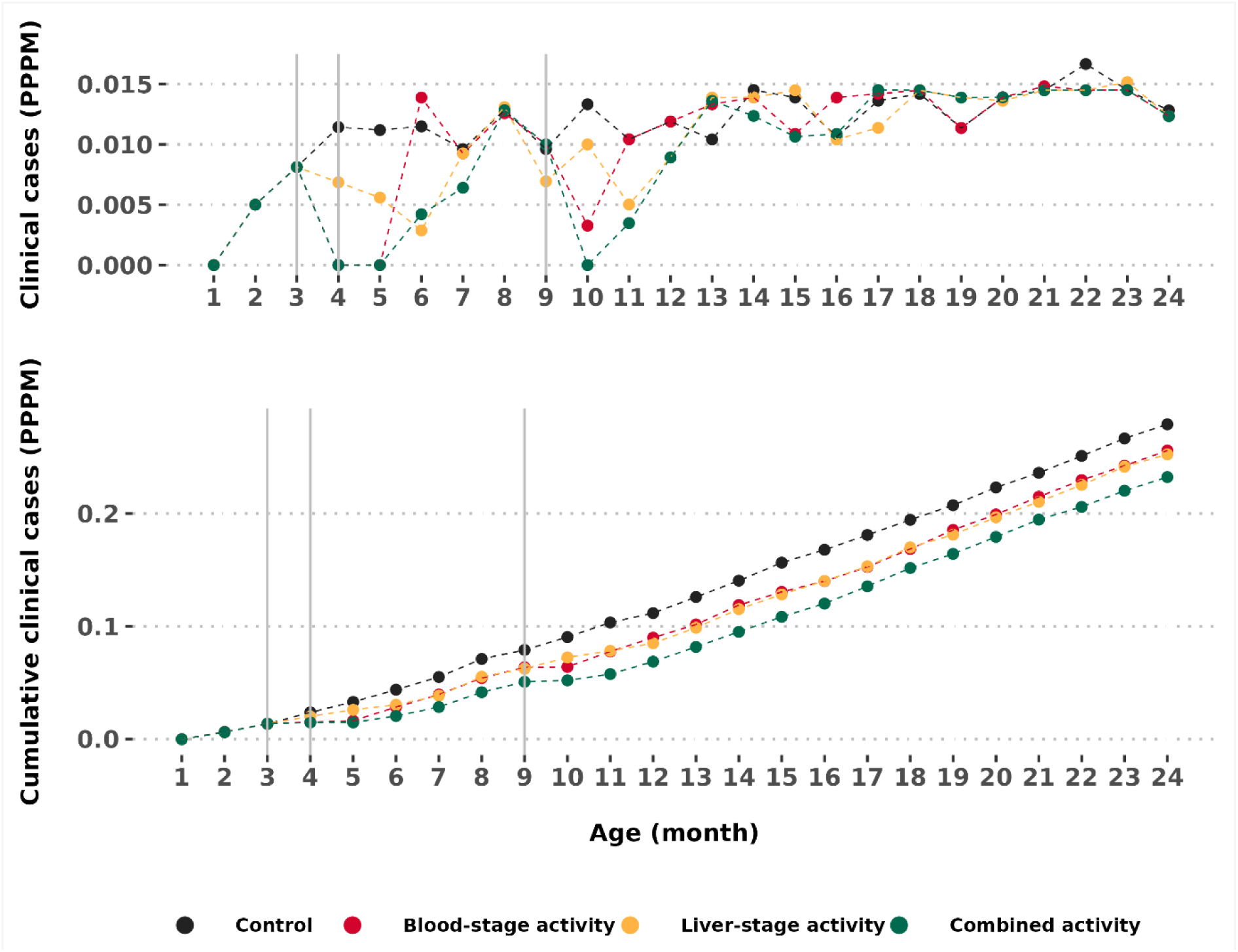
Simulated age-pattern of clinical malaria in the intervention ages in infants (up to 12 months) and post-intervention ages in the trial setting. The top panel shows the clinical incidence per age while the bottom panel shows the cumulative clinical incidences over age. The grey vertical lines indicate three IPTi-SP dosing time points. IPTi-SP: Intermittent preventive treatment in infants with sulphadoxine-pyrimethamine.

